# Single cell sequencing reveals expanded cytotoxic CD4+ T cells and two states of peripheral helper T cells in synovial fluid of ACPA+ RA patients

**DOI:** 10.1101/2021.05.28.21255902

**Authors:** Alexandra Argyriou, Marc H. Wadsworth, Adrian Lendvai, Stephen M. Christensen, Aase H. Hensvold, Christina Gerstner, Kellie Kravarik, Aaron Winkler, Vivianne Malmström, Karine Chemin

**Author notes:** **Correspondence to:** Karine Chemin, Division of Rheumatology, Department of Medicine, KI, Stockholm. Equal contribution (Alexandra Argyriou, Marc H. Wadsworth).

## Abstract

Rheumatoid arthritis is an autoimmune disease affecting the synovial joints where different subsets of CD4^+^ T cells are suspected to play a pathogenic role. So far, our understanding of the contribution of cytotoxic CD4^+^ T cells is incomplete, particularly in the context of the recently described peripheral helper T-cell subset (T_PH_). Here, using single cell sequencing and multi-parameter flow cytometry, we show that cytotoxic CD4^+^ T cells are enriched in synovial fluid of anti-citrullinated peptides antibody (ACPA)-positive RA patients. We identify two distinct T_PH_ states differentially characterized by the expression of *CXCL13* and *PRDM1*, respectively. Our data reveal that the adhesion G-Protein Coupled Receptor 56 (GPR56), a marker of circulating cytotoxic cells, delineates the synovial T_PH_ CD4^+^ T-cell subset. At the site of inflammation, GPR56^+^CD4^+^ T cells expressed the tissue-resident memory markers LAG-3, CXCR6 and CD69. Further, TCR clonality analysis revealed that most expanded clones in SF are contained within the cytotoxic and the CXCL13^+^ T_PH_ CD4^+^ T-cell populations. Finally, the detection of common TCRs between the two T_PH_ and cytotoxic CD4^+^ T-cell clusters suggest a shared differentiation. Our study provides comprehensive immunoprofiling of the heterogenous T-cell subsets at the site of inflammation in ACPA+ RA and suggests GPR56 as a therapeutic target to modulate T_PH_ cells and cytotoxic CD4^+^ T cell function.

## Introduction

CD4^+^ T cells with cytotoxic functions (CD4^+^ CTL) have gained attention in recent years and accumulating evidence supports their importance in the defense against human viral infections such as CMV^1^, EBV^2^, dengue^3^, HIV^4,5^ and SARS-CoV-2^6^. They share common functional characteristics with NK and CD8^+^ T cells, including the expression of cytolytic proteins PRF1 (perforin-1), GZMB (granzyme B)^7^ and NKG7 (Natural Killer Cell Granule Protein 7)^5^. CD4^+^ CTL can originate from Th1 cells^8^ but can also differentiate from a distinct precursor expressing the class I-restricted T cell-associated molecule (CRTAM)^9^. CD4^+^ CTL differentiation processes are still unclear but antigen dose^10^, IL-2^11^ and type 1 IFN^12^ signaling are suspected to contribute. 4-1BB (CD137) co-receptor triggering induces CD4^+^ CTL differentiation through the induction of the transcription factor Eomesodermin (Eomes)^13^ and CD4^+^ CTL cytotoxic capacity is increased after IL-15 exposure^14^. Eomes cooperates with Runx3 to induce the transcription of *Prf1* in mouse CD8^+^ T cells^15^. Together with Blimp-1, the transcription factor Hobit sustains the transcription of *Gzmb*^16^. Hobit and Blimp are also involved in the maintenance of tissue resident memory T cells (T_RM_)^17^. Both Eomes^18,19^ and Hobit^20^ are expressed in circulating human cytotoxic CTL, including CD4^+^ CTL, suggesting that they convey similar functions in a human setting. GPR56 is an adhesion G-protein coupled receptor encoded by the *ADGRG1* gene expressed on human circulating NK, CD8^+^ and CD4^+^ CTL^21^. Many of the molecular regulators of CD4^+^ CTL have been identified in mouse models or in circulating human blood. However, CD4^+^ CTL have not been investigated in great detail at the site of inflammation in human inflammatory disorders, especially in the context of T_RM_. In this environment, it is still unclear from which precursors they differentiate from.

Rheumatoid arthritis (RA), which affects around 1% of the global population, is an autoimmune disease characterized by articular bone erosion leading to physical disability, pain and decreased quality of life. Bone damage is particularly exacerbated in the subset of RA patients who present with antibodies against citrullinated proteins (ACPA)^22,23^. Although the use of biologics has revolutionized the treatment of RA, 30-40% of patients fail to respond to treatment stressing the need for novel therapeutic targets^24^. The role of CD4^+^ T cells in RA pathogenesis is evidenced by the HLA-DRB1 association with ACPA+ RA^25^. Expanded patient-specific T-cell clones have been identified in the synovium and synovial fluid (SF) of RA patients using bulk TCRβ repertoire analysis^26^. Over the years, several CD4^+^ T cell subsets, including Th1 and Th17 cells have been described in the synovial joints of RA patients^27^. Recently, PD-1^high^ HLA-Class II^+^ peripheral helper T cells (T_PH_) were identified in the synovial fluid and tissue of seropositive RA patients^28,29^ where they are proposed to facilitate B-cell recruitment and activation through CXCL13 and IL-21 production; however, there is still no satisfying marker that defines the T_PH_ subset. It is also unclear how this population is generated in the synovial joint. We have recently shown that CD4^+^ T cells with cytotoxic features are observed in SF of RA patients carrying the *PTPN22* 1858T risk allele variant^30^. This finding, as well as previous studies reporting expanded CD4^+^ CD28^null^ cytotoxic T cells in the blood of RA patients^31,32^, suggests that CD4^+^ CTL might contribute to RA. However, a transcriptomic and clonality analysis of CD4^+^ CTL, in the context of previously described CD4^+^ T cell subsets, is still missing. The identification of T-cell effector functions is deemed important since it can lead to new therapeutic strategies.

Here, we performed single cell sequencing in combination with 5′ TCRαβ sequencing on peripheral blood (PB) and synovial fluid (SF) from ACPA+ RA patients to investigate CD4^+^ CTL. We demonstrate that clonal expansions are prominent amongst CD4^+^ CTL in SF of ACPA+ RA. We also describe two T_PH_ CD4^+^ T-cell clusters differentially characterized by the expression of *PRDM1* and *CXCL13*, suggestive of two different stages of T_PH_ differentiation. Finally, we find that T_PH_ cells express the receptor GPR56 and the T_RM_ markers LAG-3, CXCR6 and CD69 implicating that these T cells are maintained in the ACPA+ RA synovial joint. Our data provide a comprehensive immunoprofiling map of pathogenic CD4^+^ T cells at the site of inflammation in ACPA+ RA.

## Results

### CD4^+^ T cells expressing cytotoxic effector molecules are enriched in synovial fluid of ACPA+ RA patients

SFMC from ACPA- (n=9) and ACPA+ (n=12) RA patients were screened for the expression of cytotoxic effector molecules and transcription factors in both CD4^+^ and CD8^+^ T cells by flow cytometry (Fig. 1, supplementary Fig. 1). ACPA+ SFMC presented with a significantly increased frequency of GZMB^+^ PRF1^+^ (p=0.0072), Hobit^+^ (p=0.0018), NKG7^high^ (p=0.0036) and GPR56^+^ (p=0.0007) CD4^+^ cells (Fig. 1a-b). No significant difference was observed for the expression of GZMA and Eomes. Amongst CD8^+^ T cells, Hobit expression was increased in ACPA+ SFMC (p=0.0170) whereas a similar trend was observed for the expression of GZMA, GZMB and NKG7 without reaching statistical significance (supplementary Fig. 2). No significant difference was observed for the expression of PRF1 and the frequency of GPR56 in CD8^+^ T cells in ACPA+ versus ACPA-RA. The frequency of GZMB^+^ PRF1^+^ CD4^+^ T cells in SF positively correlated with CCP (cyclic citrullinated peptides) antibody titers (r=0.5701, p=0.0070) (Fig. 1c). In ACPA+ synovial fluid, GPR56 identified a distinct CD4^+^ T cell population with a frequency that also correlated with CCP antibody titers (r=0.7871, p<0.0001) (Fig. 1d).

**Figure 1.**
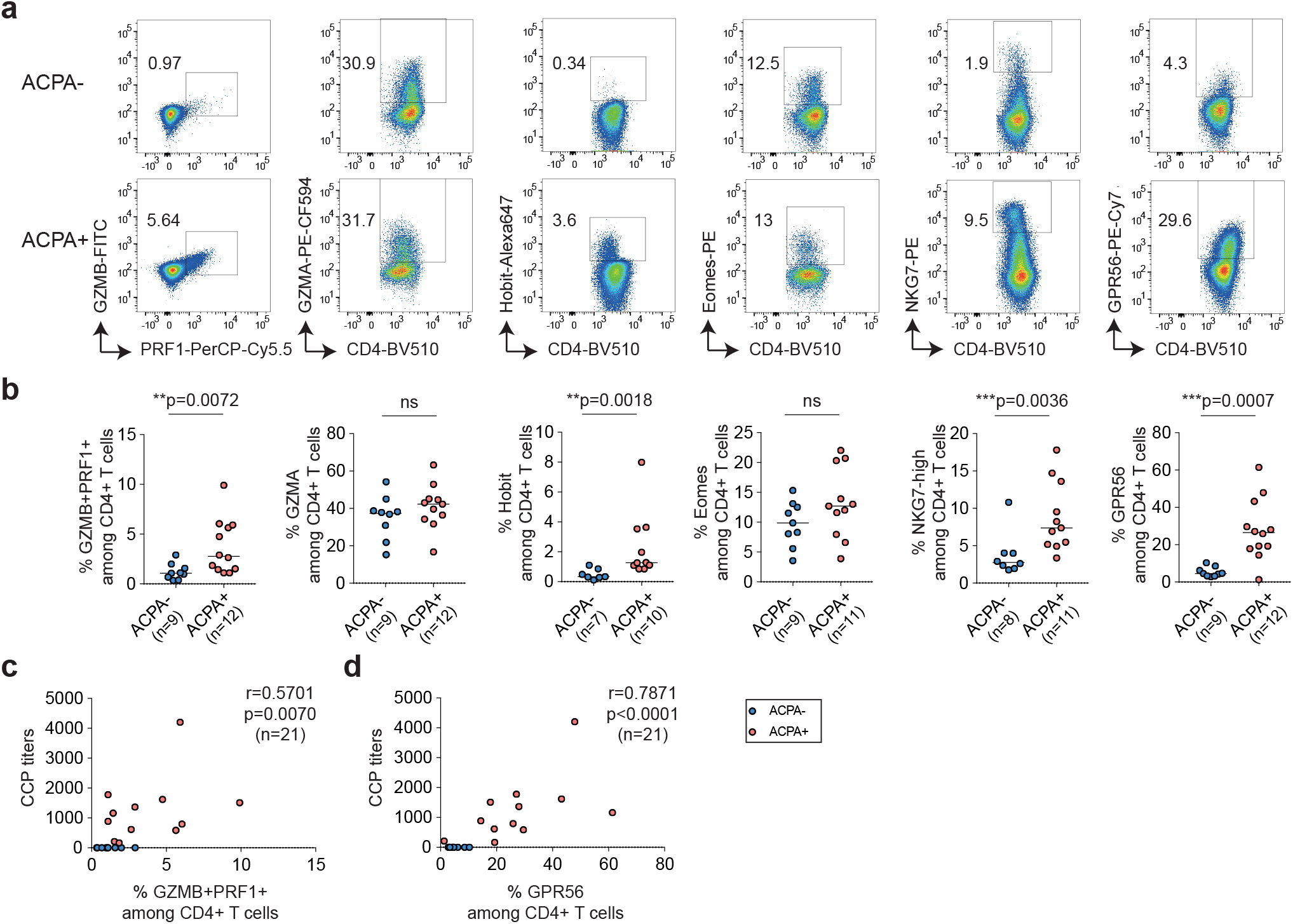
Cytotoxic CD4+ T-cell frequency in synovial fluid of ACPA- and ACPA+ RA patients. **a**) Representative flow cytometry dot plot staining of effector molecules, receptors and transcription factors associated with cytotoxic functions in CD4+ T cells from synovial fluid (SF) from ACPA-(upper panel) and ACPA+ (lower panel) RA patients, quantified in **b**) (ACPA-, n=7-9) (ACPA+, n=10-12). Line represents median, two-tailed Mann-Whitney U test, ns: not significant. **c**) Correlation between the frequency of GZMB+ PRF1+ in CD4+ T cells in SF and the level of serum anti-CCP (cyclic citrullinated peptides) antibodies levels, n=21, Spearman two-tailed test. **d**) Correlation between the frequency of GPR56 in CD4+ T cells in SF and the level of anti-CCP (cyclic citrullinated peptides) antibodies levels, n=21, Spearman two-tailed test. **a-d**) Data are from a pool of nine independent experiments where a circle is a single replicate. Blue dots indicate ACPA-RA SF and red dots indicate ACPA+ RA SF.

### Single cell sequencing identifies a cluster of cytotoxic CD4^+^ T cells and two states of T_PH_ CD4^+^ T cells in ACPA+ SFMC

To further characterize cytotoxic CD4^+^ T cells in SF of ACPA+ RA, we performed 10X single cell sequencing in combination with 5′ TCRαβ sequencing on purified CD4^+^ T cells from paired SFMC and PBMC from seven ACPA+ RA patients (Fig. 2a, supplementary table 1). After quality control (supplementary Fig. 3a), we obtained transcriptomic data from 66,360 CD4^+^ T cells in SFMC and 69,373 CD4^+^ T cells in PBMC. Unsupervised analysis of the transcriptomic data from all CD4^+^ T cells (n=135,733) from PB and SF using the Seurat package^33^ generated a total of 16 clusters annotated based on known and high scoring genes (Fig. 2b-c, supplementary Fig. 3 and 4). All 16 clusters were detected in both PB and SF from all patients although with different frequencies (Fig. 2d, supplementary table 2). A Treg cluster (cluster 15) was annotated based on the expression of *FOXP3* and *CTLA-4*^34^. Resting naïve or central memory (CM) CD4^+^ T cells express *SELL, CCR7* and *IL7R*^*35*^ (cluster 10). Two proliferating T-cell subsets expressing *MKI67* and *HELLS* were identified (clusters 13 and 14). Based on the expression of *CXCL13, MAF, TOX, PRDM1, TIGIT and PDCD-1*^*28,36,37*^, two T_PH_ cell clusters were subclassified as T_PH_ *CXCL13*^+^ (cluster 3) and T_PH_ *PRDM1*^+^ (cluster 12). Cluster 4 defines cytotoxic CD4^+^ T cells expressing cytotoxicity related genes including *NKG7, GZMH* and *PRF1*. This population was present in SF of all ACPA+ RA patients with a frequency ranging from 1.9 to 55% (Fig. 2d, supplementary Fig. 3b, supplementary table 2). In general, the frequency of T_PH_ *PRDM1*^+^, T_PH_ *CXCL13*^+^, Tregs and and cytotoxic CD4^+^ T cells was increased in SF as compared to PB (Fig. 2e, supplementary table 2). Hence, single cell transcriptomic data identify a subset of cytotoxic CD4^+^ T cells and show that the subset of T_PH_ cells is composed of two distinct clusters in ACPA+ RA patients.

**Figure 2.**
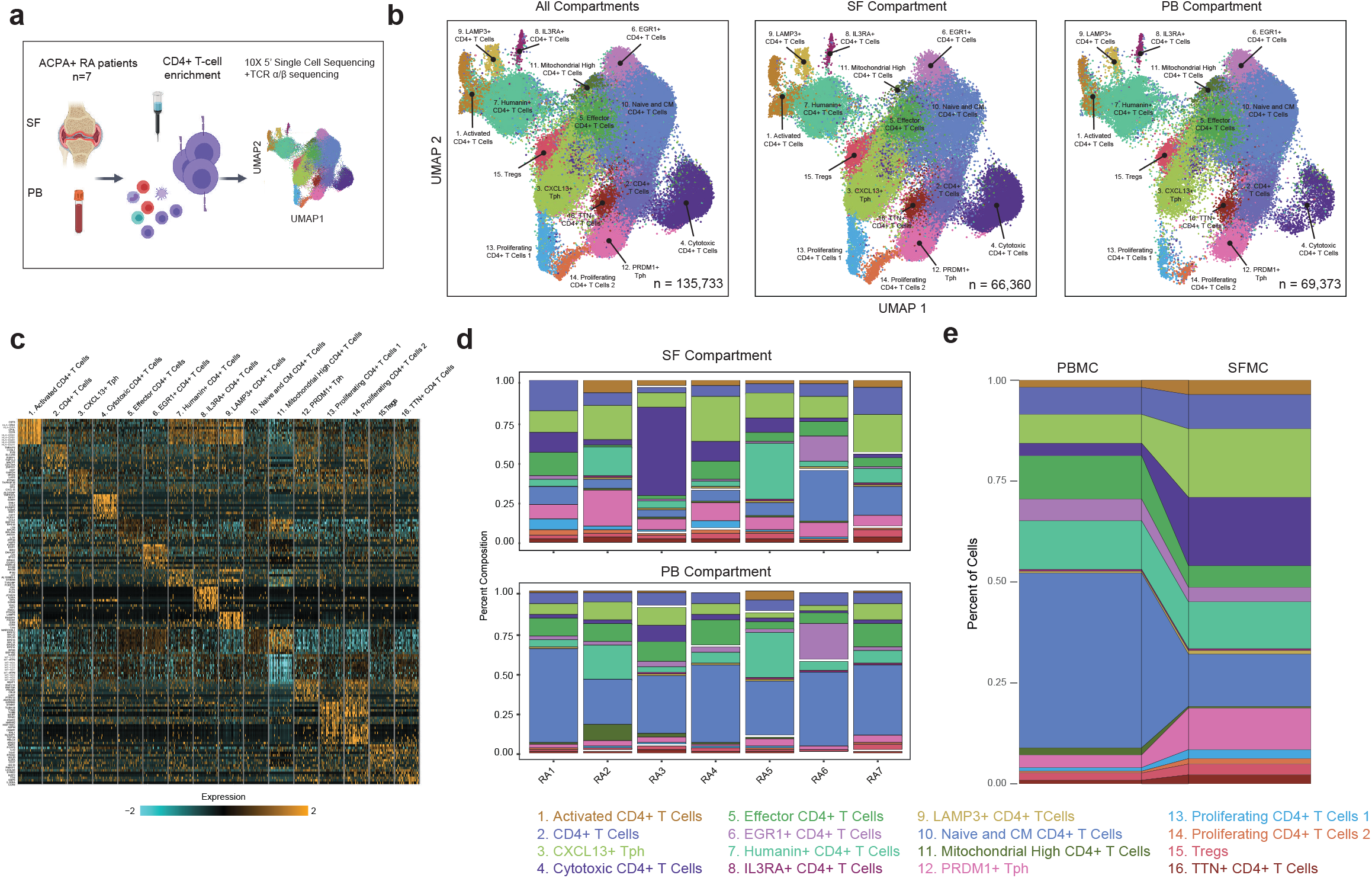
Single cell RNA sequencing of CD4+ T cells from SF and PB of ACPA+ RA patients. a) Technical workflow including CD4+ T-cell enrichment and 10X 5′single cell sequencing coupled to TCRαβ sequencing on 7 paired PB and SF from ACPA+ RA patients. **b**) UMAP plot of 135,733 cells colored by cell type cluster split by combined compartments (left), SF-only (middle) and PB-only (right panel) **c**) Heatmap showing gene expression values for a curated list of CD4+ T cell subset-defining genes **d**) Stacked barplots displaying cell type composition for each ACPA+ RA patient. **e**) Stacked barplots depicting comparative frequency of the CD4+ T-cell subsets found in PB and SF.

**Figure 3.**
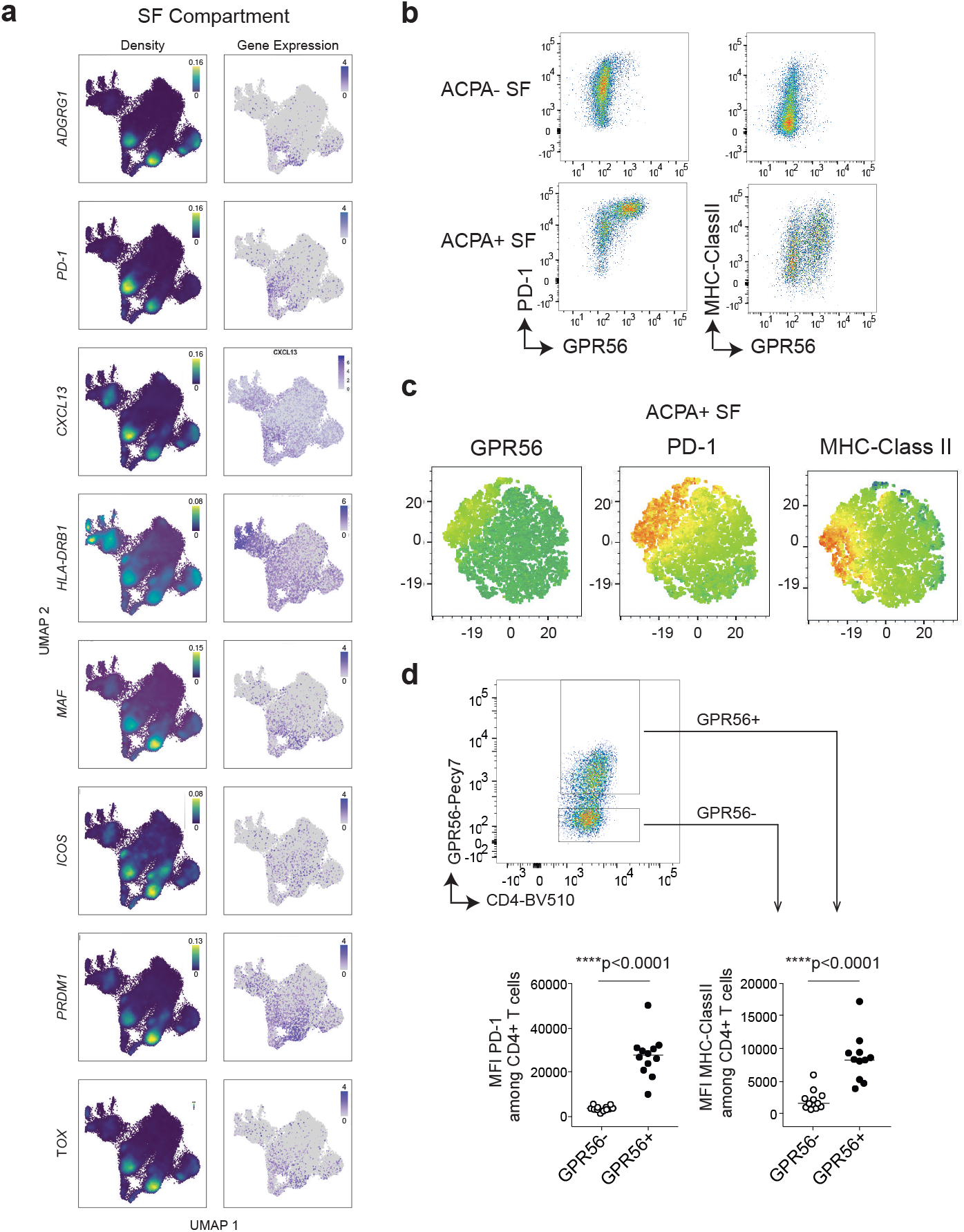
GPR56 expression on peripheral helper CD4+ T cells in ACPA+ RA SF. **a**) Nebulosa density (left panel) and gene expression (right panel) single cell plots of ADGRG1 and Tph CD4+ T-cell markers in ACPA+ RA SF. **b**) Representative flow cytometry dot plot of PD-1 and MHC-Class II staining in combination with GPR56 in ACPA- and ACPA+ RA SF. (**c**) Representative T-SNE flow cytometry plots showing CD4, GPR56, PD-1 and MHC-Class II expression in one ACPA- and one ACPA+ RA SF (gated on CD3+ T cells). (**d**) Mean fluorescence intensity (MFI) of PD-1 (n=12) and MHC-II (n=11) in GPR56-negative and positive CD4+ T cells in ACPA+ RA SF. Line represents median, two-tailed Mann-Whitney U test. Data are from a pool of nine independent experiments where a circle is a single replicate. White dots indicate GPR56-CD4+ T cells and black dots indicate GPR56+ CD4+ T cells.

**Figure 4.**
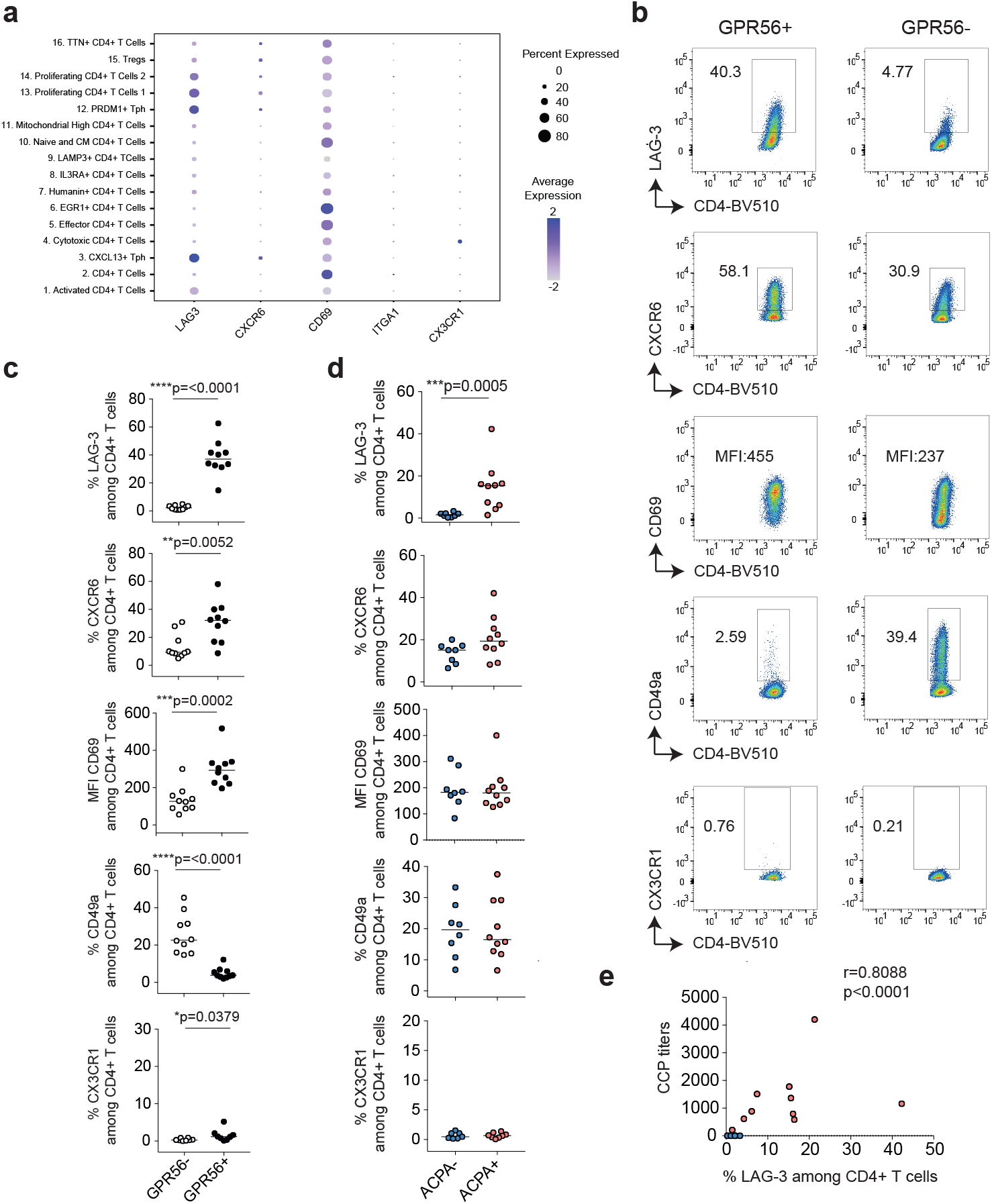
Tissue resident memory markers (T_RM_) on CD4+ T cells in ACPA+ RA SF. **a**) 2-D dot plots showing the expression of selected genes in the different CD4+ T-cell clusters in ACPA+ SF (circle size indicates cell frequency, color intensity indicates average expression). **b**) Representative flow cytometry dot plots showing the expression of LAG-3, CXCR6, CD69, CD49a and CX3CR1 on GPR56+ and GPR56-CD4+ T cells in ACPA+ RA, quantified in **c**) (n=10, LAG3, CXCR6, CD69, CD49a) (n=8, CX3CR1). White dots indicate GPR56-CD4+ T cells and blackdots indicate GPR56+ CD4+ T cells. (**d**) Expression of LAG-3,CXCR6, CD69, CD49a (n=8 ACPA-, n=10 ACPA+) and CXC3CR1 (n=7 ACPA-, n=8 ACPA+) on CD4+ T cells in SF. Blue dots indicate ACPA-RA SF and red dots indicate ACPA+ RA SF. **c-d**) Line represents median, two-tailed Mann-Whitney U test. Data are from a pool of eight independent experiments where a circle is a single replicate. (**e**) Correlation between the frequency of LAG-3 in CD4+ T cells in RA SF and the level of anti-CCP (cyclic citrullinated peptides) antibodies levels (n=18), Spearman two-tailed test.

### GPR56 delineates peripheral helper T cells in SF of ACPA+ RA patients

Since GPR56 expression was upregulated on 26% of CD4^+^ T cells in SF of ACPA+ RA patients (Fig. 1a-b), we further investigated *ADGRG1* (encoding GDPR56) in the single cell dataset. In PB, *ADGRG1* expression was expressed in T_PH_ cells (cluster 3 and 12), Humanin CD4+ T cells (cluster 7) and cytotoxic CD4+ T cells (cluster 4), the latter being characterized by the expression of *GZMA, GZMB, NKG7, PRF-1 and Hobit* (supplementary Fig. 5a-b). At the protein level, flow cytometry experiments validated that, in circulating CD4^+^ T cells, GPR56 correlated with the expression of cytotoxic effector molecules PRF1, GZMB, GZMA, NKG7 and the transcription factors Eomes and Hobit (Supplementary Fig. 5c-d) confirming a previous report^21^. Still, a smaller subset of GPR56^+^ non-cytotoxic CD4^+^ T cells could also be detected in blood (data not shown). Interestingly, in ACPA+ RA SF, *ADGRG1* expression was detected in 3 subsets: T_PH_ PRDM1 (cluster 12), T_PH_ CXCL13+ (cluster 3) and cytotoxic CD4^+^ T cells (cluster 4) with the highest expression in cluster 12 (Fig. 3a, supplementary Fig. 6). We further examined the expression of known T_PH_ associated markers^*28,36,37*^ and observed that *ADGRG1* expression overlapped with the expression of *PD-1, CXCL13, HLA-DRB1, MAF, ICOS, PDRM1* and *TOX* (Fig 3a). The two T_PH_ clusters were characterized by different levels of *ADGRG1, PD-1, CXCL13, HLA-DRB1, MAF* and *PDRM1*. Flow cytometry experiments confirmed that, in SF of ACPA+ RA patients, GPR56 expression defines the subset of T_PH_ cells (Fig. 3b-c) as illustrated by the increased expression of PD-1 (p<0.0001) and MHC-II (p<0.0001) in GPR56^+^ CD4^+^ T cells when compared to GPR56^-^ CD4^+^ T cells (Fig. 3d). GPR56 was not detected on cytotoxic CD4^+^ T cells in SF (Supplementary Fig. 6c-d) as opposed to *ADGRG1* (Fig 3a, 6a-b), highlighting the difference between transcriptomic and protein expression. These data indicate that, while GPR56 is a marker for cytotoxic CD4^+^ T cells in PB, it also delineates the subsets of PD-1^high^ MHC-Class-II^+^ T_PH_ cells in ACPA+ RA SF.

**Figure 5.**
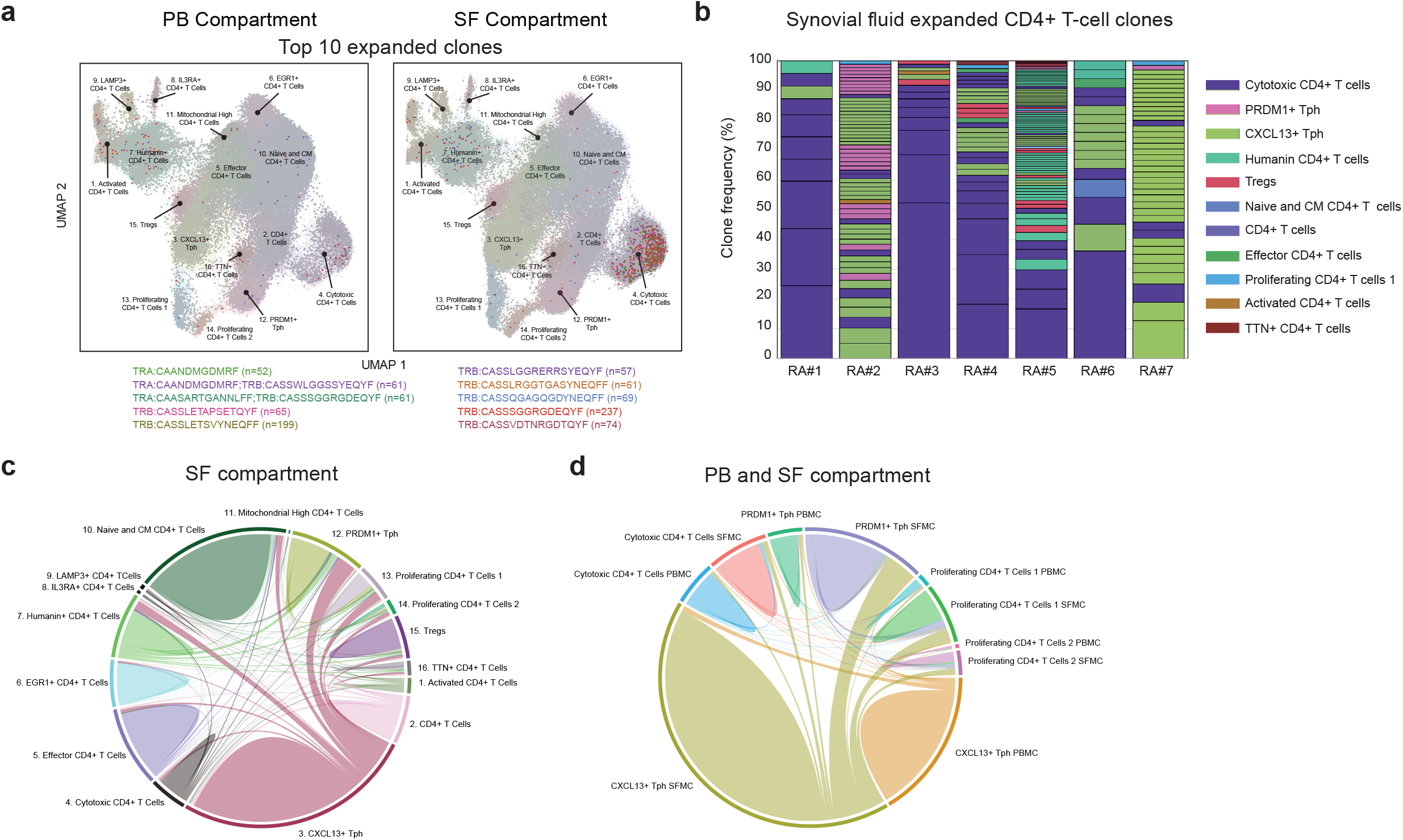
Clonally expanded CD4+ T cells in ACPA+ RA. **a**) UMAP plots showing the top 10 expanded clones in ACPA+ RA PB and SF (Each dot displays a cell). Each individual clone sharing the same CDR3 (from TCRα, TCRβ or both) amino acid sequences is highlighted by a different color. **b**) Stacked barplots displaying the phenotype of the SF expanded clones (n>4 cells) in each ACPA+ RA patient, quadrant represents individual clone. **c**) Chord diagram showing the interconnectivity between CD4+ T cells sharing the same CDR3 (from TCRα, TCRβ or both) amino acid sequenceswithin CD4+ T-cell clusters in SF of 7 ACPA+ RA patients. **d**) Chord diagram showing the interconnectivity between CD4+ T cells sharing the same CDR3 (from TCRα, TCRβ or both) amino acid sequences within selected CD4+ T-cell clusters: PDRM1+ Tph, CXCL13+ Tph, cytotoxic CD4+ T cells,proliferating CD4+ T cells in paired PB and SF.

**Figure 6.**
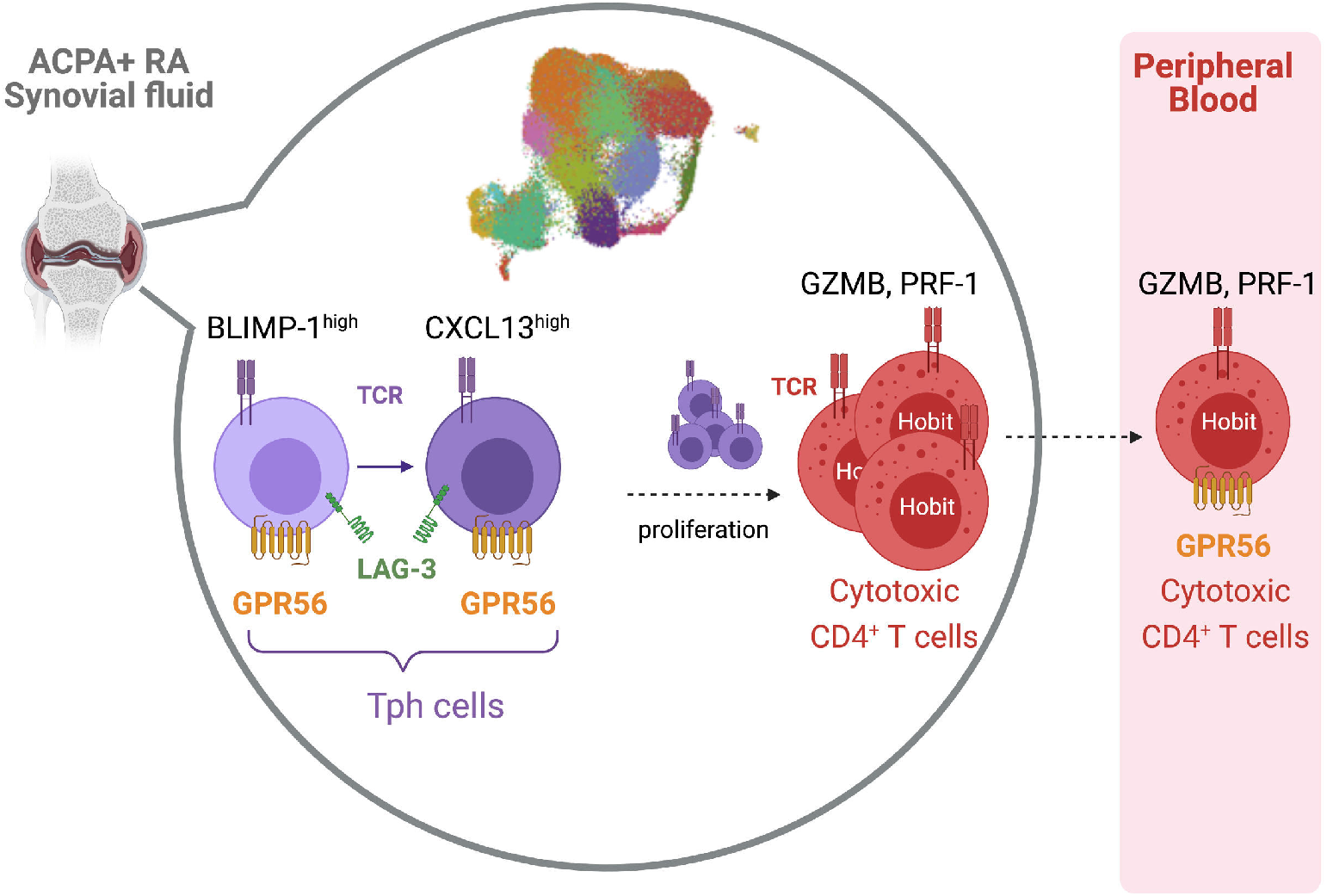
Graphical abstract showing a model for the differentiation of two states of peripheral helper T cells and cytotoxic CD4+ T cells in ACPA+ RA SF.

### GPR56^+^ CD4^+^ T cells display distinct tissue-resident memory receptors

Besides being an inhibitory receptor, PD-1 is also included in the signature associated to T-cell tissue residency^38^ and implicated in follicular helper T cells maintenance in germinal centers^39^. We therefore assessed the expression of known tissue resident memory (T_RM_) T-cell markers: *LAG3, ITGA1* (CD49a), *CD69, CXCR6* but also *CX3CR1* which is downregulated on T_RM_ T cells^38^. In SF, we found that *LAG-3* was mainly expressed on T_PH_ cells (cluster 3 and 12) and proliferating T cells (clusters 13 and 14) (Fig. 4a). A similar tendency was also observed for *CXCR6*. These genes were not expressed on cytotoxic CD4^+^ T cells, consistent with their recirculation in the blood. Since we previously observed that GPR56 delineates the subset of T_PH_ cells, we assessed the expression of tissue-resident memory markers in the context of GPR56 by flow cytometry (Supplementary Fig.7). LAG-3 (p<0.0001) and CXCR6 (p=0.0052) frequencies as well as CD69 (p=0.0002) expression was enriched on synovial GPR56^+^ CD4^+^ T cells (Fig. 4b-c). In contrast, CD49 frequency was higher on GPR56-negative CD4^+^ T cells (p<0.0001). In general, LAG-3 frequency was also significantly increased on CD4^+^ T cells in ACPA+ SF as compared to ACPA-SF (p=0.0005) (Fig. 4d) and correlated with CCP titers (r=0.8088, p<0.0001) (Fig. 4e). In CD8^+^ T cells, CD49a was the only marker for which an increased frequency was observed in ACPA+ SF (p=0.0343) (supplementary Fig. 8). These data indicate that, in ACPA+ SF, GPR56 expression correlates with the concomitant expression of a distinct set of tissue-resident memory markers on CD4^+^ T cells: LAG3, CXCR6 and CD69.

### Clonally expanded cytotoxic and CXCL13^+^ T_PH_ CD4^+^ T cells are identified in synovial fluid of ACPA+ RA patients

Next, we examined the expanded TCR clones in relation to the aforementioned CD4^+^ T-cell clusters. A clone was defined by at least 2 cells sharing the same CDR3 amino acid sequence, either from TCRα, TCRβ or the combination of both. The 10 most expanded clones were primarily identified in the cytotoxic CD4^+^ T-cell subset and to a lesser extent in the Humanin^+^ CD4^+^ T-cell subset in ACPA+ RA SF (Fig. 5a, right panel and supplementary Table 3 and 4). Some of the expanded clones were shared between PB and SF (Fig. 5a, left panel). These 10 expanded T-cell clones were originating from 4 RA patients (Supplementary Figure 9) of which three identical clones were identified between patient 5 and 6. The analysis of synovial fluid T-cell clones composed of at least 4 cells in each patient revealed that they mainly originate from cytotoxic CD4^+^ T cells, CXCL13^+^ T_PH_ and to a lesser extent PRDM1^+^ T_PH_ and Humanin CD4^+^ T cells (Fig. 5b, supplementary Table 3 and 4). Further, we examined overlapping CDR3 sequences between SF CD4^+^ T-cell subsets that would be indicative of a possible shared differentiation pathway (Fig. 5c). In SF, the CXCL13^+^ T_PH_ cluster shared TCR sequences with the PRDM1^+^ T_PH_, Humanin^+^ CD4^+^ and proliferating CD4^+^ T-cell clusters. Common CDR3s were also identified between cytotoxic CD4^+^ T cells and *CXCL13*^+^ T_PH_ cells. Of note, in SF, Tregs shared only few CDR3 sequences with other CD4^+^ T-cell subsets, suggesting that they could be thymus-derived Tregs^40^ and not induced at the site of inflammation from conventional CD4^+^ T cells. We then further focused our TCR analysis on the clusters of interest (cytotoxic CD4^+^ T cells, PRDM1^+^ T_PH_, CXCL13^+^ T_PH_ and proliferating CD4^+^ T cells 1 and 2) in PB and SF (Fig. 5d). We observed that some CDR3 sequences were shared between cytotoxic CD4^+^ T cells in PB and SF. Similarly, CXCL13^+^ T_PH_ cells had common CDR3s in PB and SF. Overall, these data show that, in ACPA+ synovial fluid, expanded clones are mainly found within the cytotoxic and the CXCL13^+^ T_PH_ CD4^+^ T-cell subsets and that shared CDR3 sequences are identified between clusters in particular between the two T_PH_ clusters and between CXCL13^+^ T_PH_ and cytotoxic CD4^+^ T cells.

## Discussion

Cytotoxic CD4^+^ CD28^null^ T cells are expanded in the blood of patients with RA^31^ but their presence and clonality at the site of inflammation in ACPA+ RA was still not deeply characterized. Leveraging single cell technology, we identified the presence of cytotoxic CD4^+^ T cells in ACPA+ RA SF that are clonally expanded in synovial fluid and, for some patients, in their corresponding peripheral blood. We further define the T_PH_ CD4+ T-cell subset at the single cell level showing that it is composed of two independent clusters with different *CXCL13* and *PRDM1* (Blimp-1) expression levels. So far, the T_PH_ CD4^+^ T-cell subset was only defined by high expression of PD-1 and MHC-Class-II and more accurate markers were still lacking. We show that GPR56 clearly delineates the T_PH_ subset and associates with the expression of tissue resident memory markers LAG-3, CXCR6 and CD69. Together, these data provide a refined characterization of cytotoxic and T_PH_ CD4^+^ T cells, two pathogenic CD4^+^ T-cell subsets in ACPA+ RA (Fig. 6).

In synovial tissue of ACPA+ RA patients, T_PH_ CD4^+^ cells produce CXCL13^28,36,41^, localize with B cells and have the ability to induce *in vitro* plasma cell differentiation through IL-21 production^28^. T_PH_ cells are defined by the high expression of PD-1 and MHC-Class II, the cytokines CXCL13, IL-21 and IL-10, and the transcription factors Blimp-1, TOX, MAF and SOX4^*28,36,37*^. T_PH_ CD4^+^ T-cell differentiation is induced by TGFβ in IL-2 neutralizing conditions^37^. Single cell sequencing identified two distinct clusters of T_PH_ cells: PRDM1^+^ T_PH_ cells and CXCL13+ T_PH_ cells. We postulate that these two clusters correspond to two states of differentiation: PRDM1^+^ T_PH_ CD4^+^ T cells could be the precursors of the more differentiated cytokine producing CXCL13^+^ T_PH_ CD4^+^ T cells. Indeed, Blimp-1 encoded by *PRDM1* negatively regulates the expression of CXCL13^37^. In line with this hypothesis, common TCR sequences observed between PRDM1^+^ T_PH_ cells and CXCL13^+^ T_PH_ cells clusters suggest of a common origin between these two subsets.

The impact of CD4^+^ CTL in the synovial joint is less well characterized. Expansions of CD4^+^ CTL are frequently observed in chronic viral infections and CD4^+^ CTL are capable of directly killing EBV-infected B cells in a MHC-class II dependent manner^2^. Whether CD4^+^ CTL are protective or pathogenic in viral infections is still a matter of debate and their role may vary depending on the type of infections^5^. Recently, single cell sequencing data has also identified infiltrating CD4^+^ CTL in liver cancer^42^ and in bladder cancer where they provided anti-tumor killing in the context of MHC-Class II^43^. Interestingly, a CD4^+^ CTL signature in bladder tumors predicted clinical response to anti-PD-L1^43^. Furthermore, expansions of cytotoxic CD28^null^ CD4^+^ T cells have been repeatedly reported in the blood of patients with rheumatic diseases such as RA^31^, myositis^44^ and vasculitis^45^ as well as in cardiovascular diseases^46,47^. PRF1^+^ CD4^+^ T cells were also identified by immunofluorescence in synovial tissue of RA patients^48^. Our data shows that cytotoxic CD4^+^ T cells are enriched in SF of ACPA+ RA patients and correlates with CCP titers showing that cytotoxic CD4^+^ T cells are mainly implicated in the subset of autoantibody-positive RA. Purified perforin-1^49^ as well as supernatants from IL-15 stimulated synovial CD8^+^ T cells^50^ induces histone citrullination in neutrophils suggesting that cytotoxic effector molecules and citrullination are tightly linked in RA. In our dataset, cytotoxic CD8^+^ T cells also tended to be more cytotoxic in ACPA+ RA and could contribute to a perforin-dependent citrullination mechanism. The presence of CD4^+^ CTL only in SF of ACPA+ RA suggest that they have a pathogenic function, but further studies are warranted to understand the functional consequences of their interactions with antigen presenting cells in the synovial joint.

The differentiation processes that lead to the differentiation of cytotoxic CD4^+^ T cells are not completely elucidated. IL-2, IL-15 and type I IFNs, which have been implicated in the differentiation of cytotoxic CD4^+^ T cells, are also expressed in the synovial joint possibly contributing to their development. We show that the transcription factor Hobit, which has been shown to regulate *Gzmb* expression in mouse^16^, was expressed in GZMB/PRF-1-expressing CD4^+^ T cells in both PB and SF suggesting that Hobit also drive CD4^+^ CTL differentiation in ACPA+ RA. We have previously shown that PRF-1-expressing Eomes^+^ CD4^+^ T cells are increased in SF of *PTPN22* risk carriers^30^. In dengue viral infections, an increased frequency of CD4^+^ CTL was observed in HLA-DRB1*0401+ individuals^51^. Hence, RA-associated genetic polymorphisms might contribute to increase the differentiation of CD4^+^ CTL through mechanisms that remain to be investigated.

Expanded TCR clones have been identified in synovial tissues and fluid from RA patients using bulk TCRβ sequencing^26^. In blood, expanded CD4^+^ CD8^null^ cells present with a bias in their TCRVβ usage^52,53^. In our dataset, the most expanded T-cell clones identified in SF of ACPA+ RA presented with cytotoxic or CXCL13^+^ T_PH_ signatures. The differentiation towards the cytotoxic CD4^+^ T cell lineage has been proposed to be linked to repeated antigen stimulation as illustrated in the case of chronic viral infections such as CMV^10^. Expanded cytotoxic CD4^+^ T cell clones in SF might therefore be the final stage of differentiation of autoreactive CD4^+^ T cells which have been exposed to several rounds of antigen stimulation. Several studies have shown that CMV reactivity is found in the subset of CD4^+^ CD8^null^ cell subset but so far, their autoreactive properties have not been investigated in the context of citrulline immunity. In our data set, some TCR sequences were shared between T_PH_ cells and cytotoxic CD4^+^ T cells and we can speculate that T_PH_ cells, after repeated antigenic stimulation led to cytotoxic CD4^+^ T cell expanded clones.

GPR56 is an adhesion G-protein coupled receptor encoded by the *ADGRG1* gene implicated in numerous migration/adhesion processes including neuron migration^54^ and tumor growth inhibition^55^. In NK cells, GPR56 is an inhibitory receptor binding to the tetraspanin CD81^56^. We confirmed a previous report showing that GPR56 is also a marker of circulating cytotoxic CD4^+^ cells^21^. At the site of inflammation, however, we discovered that GPR56 was highly expressed on T_PH_ cells in ACPA+ RA patients and could be used as a new marker to identify T_PH_ CD4^+^ T cells. In SF CD4^+^ T cells, GPR56 expression correlated with the expression of PD-1, LAG-3, CXCR6 and CD69 expressed on tissue-resident memory T cells^38^. Similarly, in multiple sclerosis, CD8^+^ T cells from active lesions exhibit a tissue resident memory phenotype (CD69, CD103, PD-1, CD49a) associated with an intermediate expression of GPR56 without granzyme B expression^57^. In mouse, Blimp-1 and Hobit mediates the transcriptional program associated with T-cell tissue residency^17^. In ACPA+ SF, Blimp-1 was expressed on T_PH_ CD4^+^ cells presenting with tissue resident memory markers whereas Hobit was only expressed on cytotoxic CD4^+^ T cells suggesting that, Blimp-1 could drive tissue residency of T_PH_ CD4^+^ T cells in the synovial joint. In human NK cells, Hobit triggers *ADGRG1* expression^56^ but, in SF, *Hobit* was not expressed in *ADGRG*1+ CD4^+^ T cells suggesting that a different transcription factor could regulate GPR56. GPR56 might be implicated in the migration and/or maintenance of T_PH_ CD4^+^ T cells in the synovial joints where its ligand still remains to be determined.

In summary, we identify clonally expanded cytotoxic CD4^+^ T cells and two clusters of T_PH_ CD4^+^ T cells in ACPA+ SF. We show that GPR56 is a marker that should be used to identify T_PH_ CD4^+^ cells. Future studies aiming at understanding the function of GPR56 in T_PH_ CD4^+^ and cytotoxic CD4^+^ T cells will contribute to evaluate the feasibility of targeting this receptor in ACPA+ RA.

## Methods

Synovial fluid from ACPA-(n=9) and ACPA+ (n=12) RA patients were collected at the Rheumatology Clinic of Karolinska University Hospital, Stockholm (Supplementary Table 1). In n=8 ACPA- and n=8 ACPA+ RA patients, paired PBMCs were also available. All patients had established disease, based on the 1987 criteria from the American College of Rheumatology. This study was approved by the research ethics committee of Karolinska University Hospital and all patients signed informed consent according to the Declaration of Helsinki. PBMC and SFMC were isolated by Ficoll separation (GE Healthcare) and cryopreserved in fetal bovine serum (FBS) with 10% DMSO in liquid nitrogen.

### Flow Cytometry

1.10^6^ PBMC and 1.10^6^ SFMC were labeled with LIVE DEAD Fixable NEAR IR Dead Cell Stain Kit (Invitrogen) and further stained with fluorescently labeled antibodies (Supplementary table 5). ACPA- and ACPA+ SF and PB were always run in the same experiment. Depending on the number of recovered cells, 2 to 4 flow cytometry panels were run (Supplementary table 6). For intracellular stainings, cells were fixed and permeabilized using the FOXP3 permeabilization buffer kit (ebioscience) according to the manufacturer′s protocol. Samples were run on the BD LSR Fortessa and flow cytometry data was analyzed with the FlowJo software (version X 10.07r2).

### Statistical Analysis

Flow cytometry results were analyzed with the Prism 7 software (GraphPad, San Diego, CA, USA) using two-tailed Mann-Whitney U test. Statistical tests performed are indicated in the figure legends.

### Single Cell Processing

CD4^+^ T cells were enriched from cryopreserved PBMC and SFMC samples using the EasySep human CD4 Positive Selection Kit II (STEMCELL Technologies, Catalog# 17852). Following CD4^+^ T-cell isolation, cells were counted, hashed with TotalSeq-C0251 (ref: 394661) and TotalSeq-C0253 (ref: 394665) anti-human antibodies (Biolegend) and pooled per patient (i.e., pool 1 = PB and SF from patient 1, pool 2 = PB and SF from patient 2, etc.). The pooled cells were then counted again and immediately loaded onto the chromium chip G using the standard protocol for the Chromium single cell 5’ kit v1.1 (10x Genomics, Inc). Following Gel Bead-in Emulsion (GEM) generation, samples were processed using the standard manufacturer’s protocol. Once sequencing libraries passed standard quality control metrics, the libraries were sequenced on Illumina NextSeq500/550 high output 75-cycle v2.5 kits with the following read structure: read1: 26, read2: 58, index 1: 8. Libraries were sequenced to obtain a read depth greater than 20,000 reads/cell for the gene-expression (GEX) libraries and greater than 5,000 reads/cell for the V(D)J-enriched T Cell libraries.

### Alignment and Preprocessing

Following sequencing, the pooled GEX libraries were demultiplexed using bc2fastq (v2.17) with modified parameters (--minimum-trimmed-read-length=10 --mask-short-adapter-reads=10 --ignore-missing-positions --ignore-missing-controls) to generate FASTQ files for each patient. FASTQ files were then uploaded to Terra (www.app.terr.bio) where the raw sequencing data were mapped and quantified using STAR within the 10X Genomics Inc software package CellRanger on Cumulus (https://cumulus.readthedocs.io/en/latest/cellranger.html, snapshot 15, Cellranger 5.0.1, default parameters) with GRCh38-2020-A (https://cumulus.readthedocs.io/en/latest/cellranger.html) to generate UMI-collapsed, gene-by-cell expression matrices. Aligned matrices were first filtered to remove low-quality barcodes, keeping only those with greater than 200 UMIs, less than 4,000 UMIs, and less than 20% mitochondrial reads. Cell doublets were then computationally removed using DoubletFinder^58^ with a calculated doublet rate of 14% and default parameters.

### Unsupervised Analysis and Cell Type Annotation

Using the Seurat^59^ (v4.0.0) package, filtered gene-by-cell matrices were merged and then processed using a standard unsupervised workflow (i.e., normalization, scaling, dimensionality reduction, batch correction, cell clustering, and differential gene expression analysis). First, the merged matrix was normalized and log-transformed (scaling factor = 10,000). The top 3000 highly variable genes were identified and used for scaling the data. During scaling, two sources of unwanted variation, cumulative UMI capture and percent mitochondrial reads, were regressed out. Dimensionality reduction was then performed with principal component analysis (PCA) over the top 3000 variable genes and, using the JackStraw function in Seurat, we selected 53 statistically significant principal components (PCs, p < 0.01). Using these significant PCs, we corrected for compartment and patient-specific batch effects with the harmony^60^ package, and then proceeded with constructing the nearest neighbor graph and Uniform Manifold Approximation and Projection (UMAP) plots. Cells were clustered using Louvain clustering and the package clustree^61^ was used to generate a clustering tree and identify which resolution achieved stability. Differential gene expression analysis was then performed using the FindAllMarkers Seurat function with test.use set to ‘MAST’^62^. We also used Nebulsosa^63^ to visualize the joint density approximations of known T-cell phenotypic markers. We attempted to deconvolute the multiplexed PBMC and SFMC cell populations using the cell hashing antibodies, however, due to sub-optimal tagging we were unable to separate the cell populations. To address this issue, we generated a compartment-specific reference using PBMC and SFMC samples from two more volunteers and then reference-mapped our mixed samples to assign compartment membership^59^. Only barcodes with a predicted id score greater than 0.5 were kept and used for downstream analyses. Finally, non-T cells were filtered out by removing barcodes with zero gene expression for CD3 genes. Taking both the results of the differential expression analysis and joint density approximations of known phenotypic markers, we identified 16 unique T cell subsets: 1. Activated CD4^+^ T Cells (CD38^+^ HLA-DRA^+^); 2. CD4+ T Cells (IL7R^+^); 3. CXCL13^+^ Tph (CXCL13^+^ CD3D+); 4. Cytotoxic CD4^+^ T Cells (GNLY^+^ GMZB^+^ PRF1^+^ GZMA^+^ NKG7^+^); 5. Effector CD4+ T Cells (CCR6^+^ KLRB1^+^ CD27^+^); 6. EGR1^+^ CD4^+^ T Cells (EGR1^+^); 7. Humanin^+^ CD4^+^ T Cells (MTRNR2L12^+^ MTRNR2L8^+^); 8. IL3RA^+^ CD4^+^ T Cells (IL3RA^+^); 9. LAMP3^+^ CD4^+^ T Cells (LAMP3^+^); 10. Naïve and Central Memory (CM) T Cells (IL7R^+^, CCR7^+^, SELL^+^); 11. Mitochondrial High CD4^+^ T cells (MT-CO1^+^MT-ATP8^+^MT-ND4^+^); 12. PRDM1^+^ Tph (PRDM1^+^ ADGRG1^+^); 13. Proliferating CD4^+^ T Cells 1 (STMN1^+^); 14. Proliferating CD4^+^ T Cells 2 (KI67^+^); 15.Tregs (TIGIT^+^ FOXP3^+^ IL2RA^+^); 16. TTN^+^ CD4^+^ T cells (TTN^+^).

### TCR Mapping

Following sequencing, the V(D)J pooled libraries were demultiplexed using bc2fastq (v2.17.1.14) to generate FASTQ files for each patient, uploaded to terra, and mapped and quantified using the 10X Genomics Inc software package CellRanger on Cumulus (https://cumulus.readthedocs.io/en/latest/cellranger.html, snapshot 15, Cellranger 5.0.1, default parameters) to generate consensus annotation files for each patient. Using the scRepertoire^64^ package, patient-specific consensus annotation files were consolidated into a list of TCR sequencing results and then integrated with the Seurat object using the combineExpression function. Clonotypes were called based on the CDR3 gene and clonotype expansion were assigned based on the absolute frequency. Finally, the interconnectivity between specific cell types, both within and between tissue compartments, were visualized using chord diagrams.

## Data Availability

Data is available upon request.

## Acknowledgments

We thank the patients who donated samples and the medical staff at the Rheumatology Clinic of Karolinska University Hospital. Julia Boström, Gloria Rostvall, and Susana Hernandez Machado are acknowledged for organizing the sampling, storage, and administration of biomaterial. We thank Leonid Padyukov and Barbro Larsson for HLA-DR genotyping of the samples and Lena Israelsson for technical assistance with ACPA measurement. This study is supported by grants from Dr. Margaretha Nilssons, the Nanna Svartz, the Börje Dahlin and the Ulla and Gustaf af Ugglas foundations as well as the Swedish association against rheumatism. This project has received funding from the Innovative Medicines Initiative 2 Joint Undertaking (JU) under grant agreement No 777537 (RTCure). The JU receives support from the European Union’s Horizon 2020 research and innovation programme and EFPIA.

## Author contributions

A.A., A.L. and C.G. performed and analysed the flow cytometry experiments. M.H.W performed the 10X experiments and led the single cell analysis. S.M.C. performed preliminary 10X experiments and single cell data analysis. A.H.H. diagnosed RA patients and collected clinical information. K.K. and A.W. contributed to the design of the single cell experiments. V.M. contributed to the design of the study and the RA sample collection. A.A., M.H.W and KC. generated the figures. K.C designed the study, oversaw the analyses and wrote the paper. All authors read, edited and approved the paper.

