## Supplementary Figures and Tables for "Single cell sequencing reveals expanded cytotoxic CD4+ T cells and two states of peripheral helper T cells in synovial fluid of ACPA+ RA patients"

### Supplementary Fig. 1

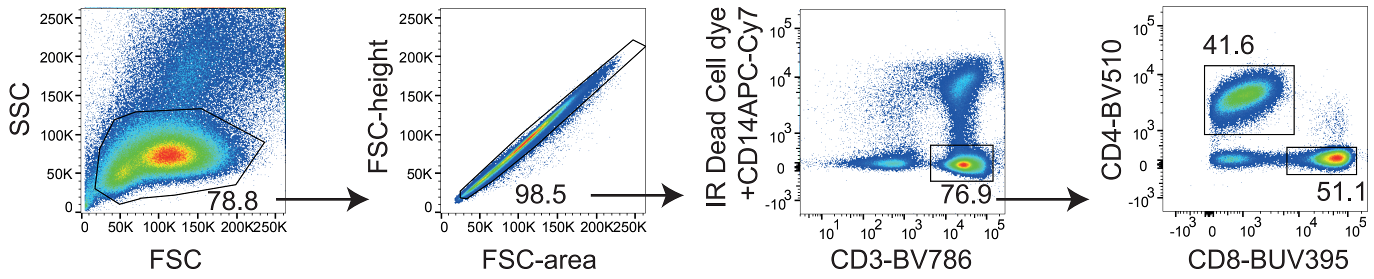


**Supplementary Fig. 1.** Facs gating strategy for Figure 1 included lymphocyte gating (based on forward and side scatter) followed by doublets, CD14+ and dead cells exclusion. CD4+ or CD8+ T cells were further selected among CD3+ T cells.

#### Supplementary Fig. 2

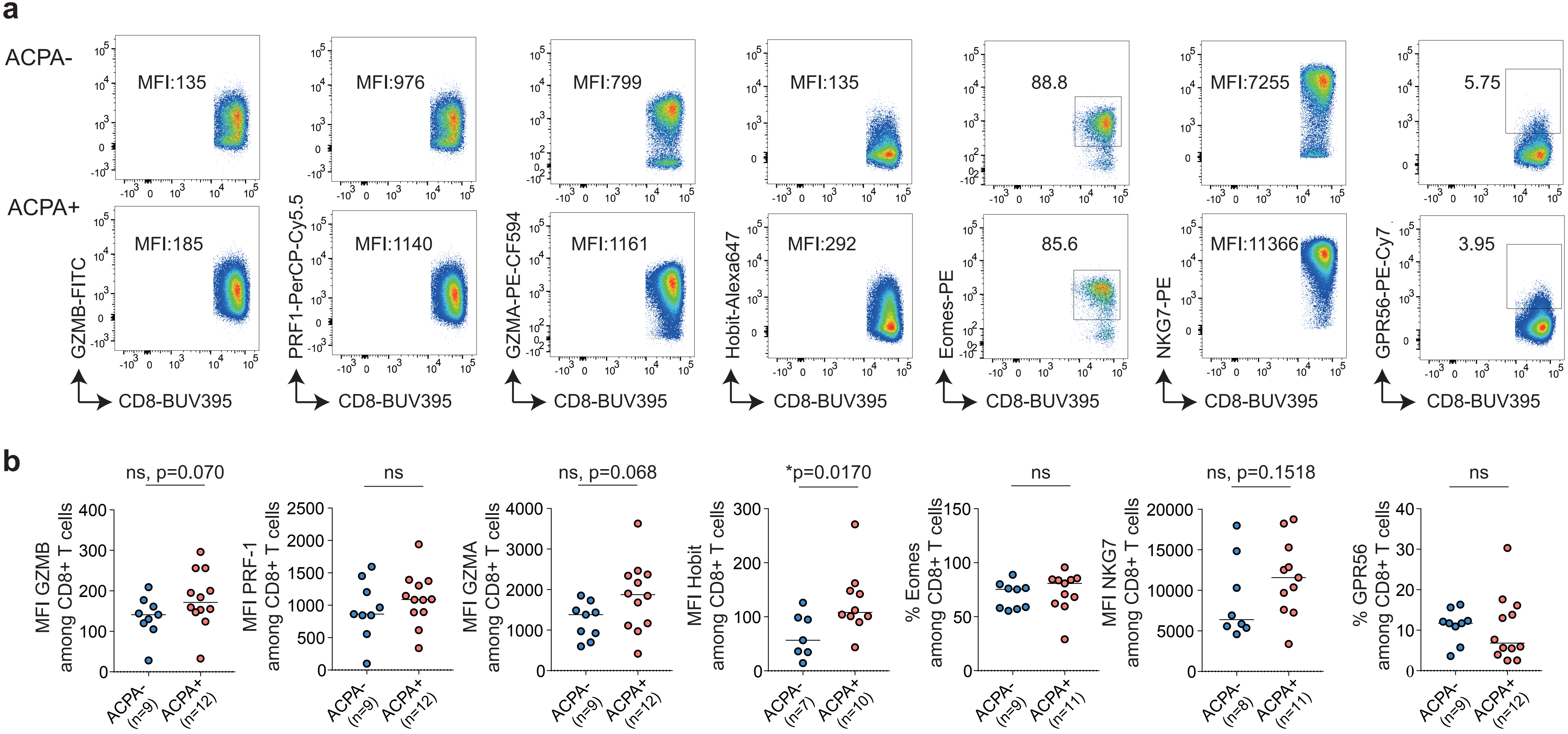


**Supplementary Fig. 2. Cytotoxic markers in CD8+ T cells in SF of RA patients**. **a**) Representative flow cytometry dot plot staining of effector molecules, receptors and transcription factors associated with cytotoxic functions in CD8+ T cells from synovial fluid (SF) from ACPA- (upper panel) and ACPA+ (lower panel) RA patients quantified in **b**), (ACPA-, n=7-9) (ACPA+, n=10-12). Line represents median, Mann-Whitney U test, ns: not significant. Data are from a pool of nine independent experiments where a circle is a single replicate. Blue dots indicate ACPA- RA SF and red dots indicate ACPA+ RA SF.

### Supplementary Fig. 3

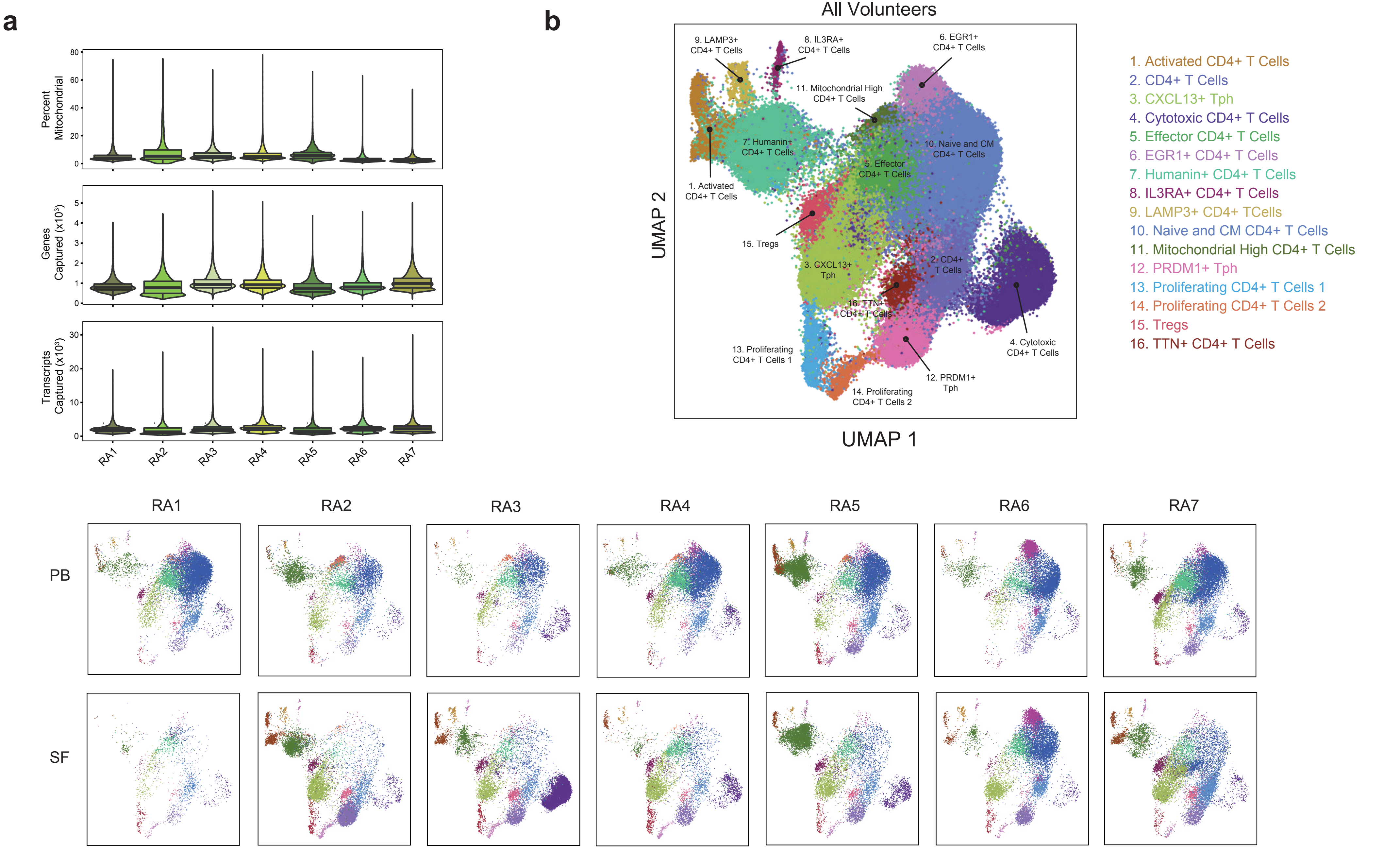


**Supplementary Fig 3. Single cell RNA sequencing of CD4+ T cells from SF and PB of ACPA+ RA patients**. a) Violin plots showing the frequency of mitochondrial genes (upper panel), the number of genes captured (middle panel) and the total number of transcripts (lower panel) per patient.
b) UMAP plot of 135,733 cells colored by cell type cluster split by patients and compartments: PB-only (upper panel) and SF-only (lower panel).

### Supplementary. Fig. 4

**a**

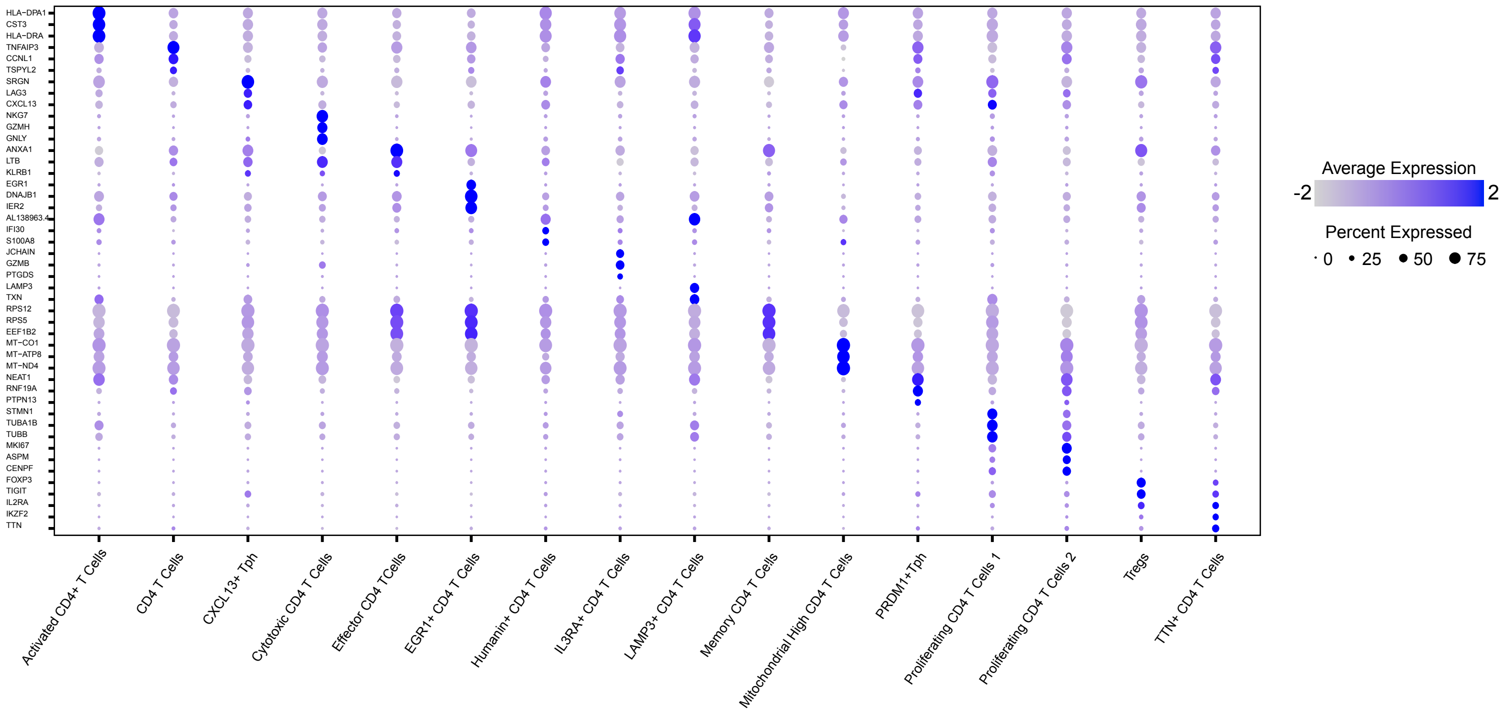


**b**

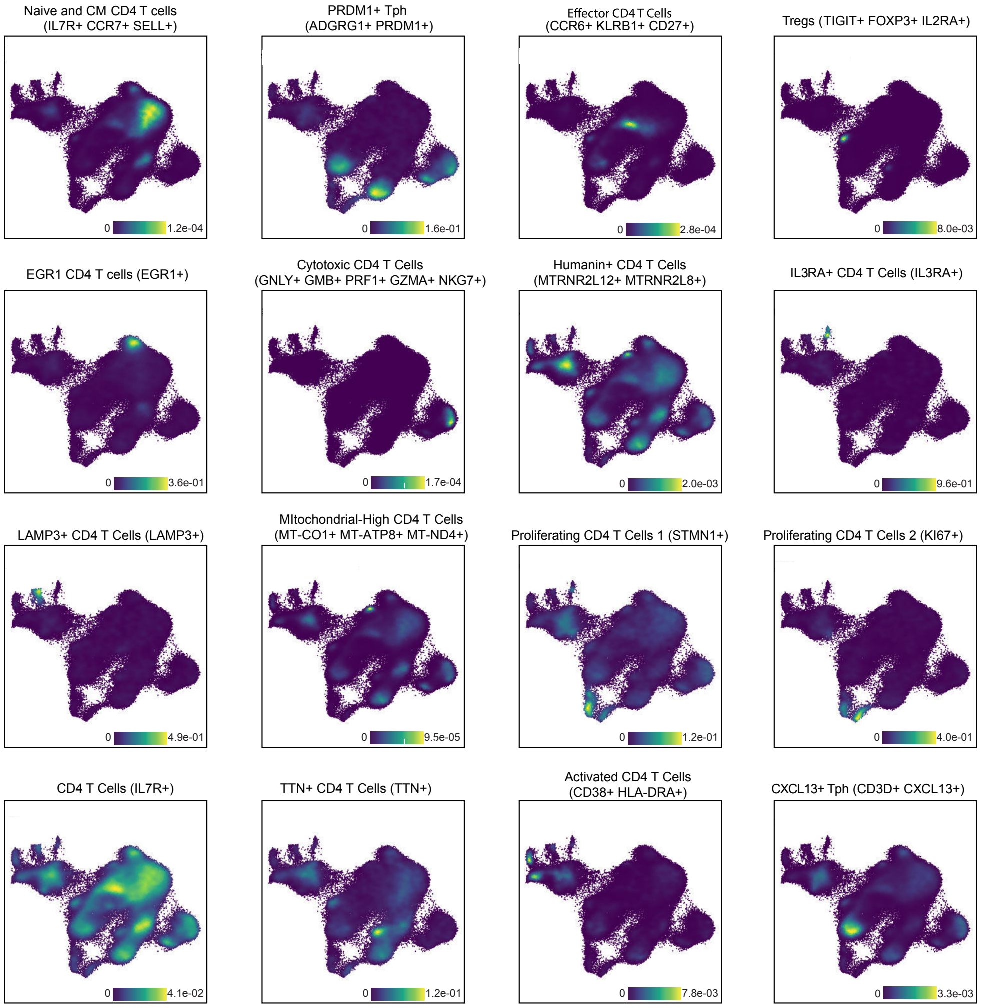


**Supplementary Fig. 4. Differentially expressed (DE) genes in CD4+ T-cell clusters in combined PB and SF single cell data from 7 ACPA+ RA patients.** a) 2-D dot plots showing the expression of the top 3 DE genes in each cluster (circle size indicates cell frequency, color intensity indicates average expression) b) Nebulosa density plots showing key genes used to annotate the clusters.

### Supplementary Fig. 5

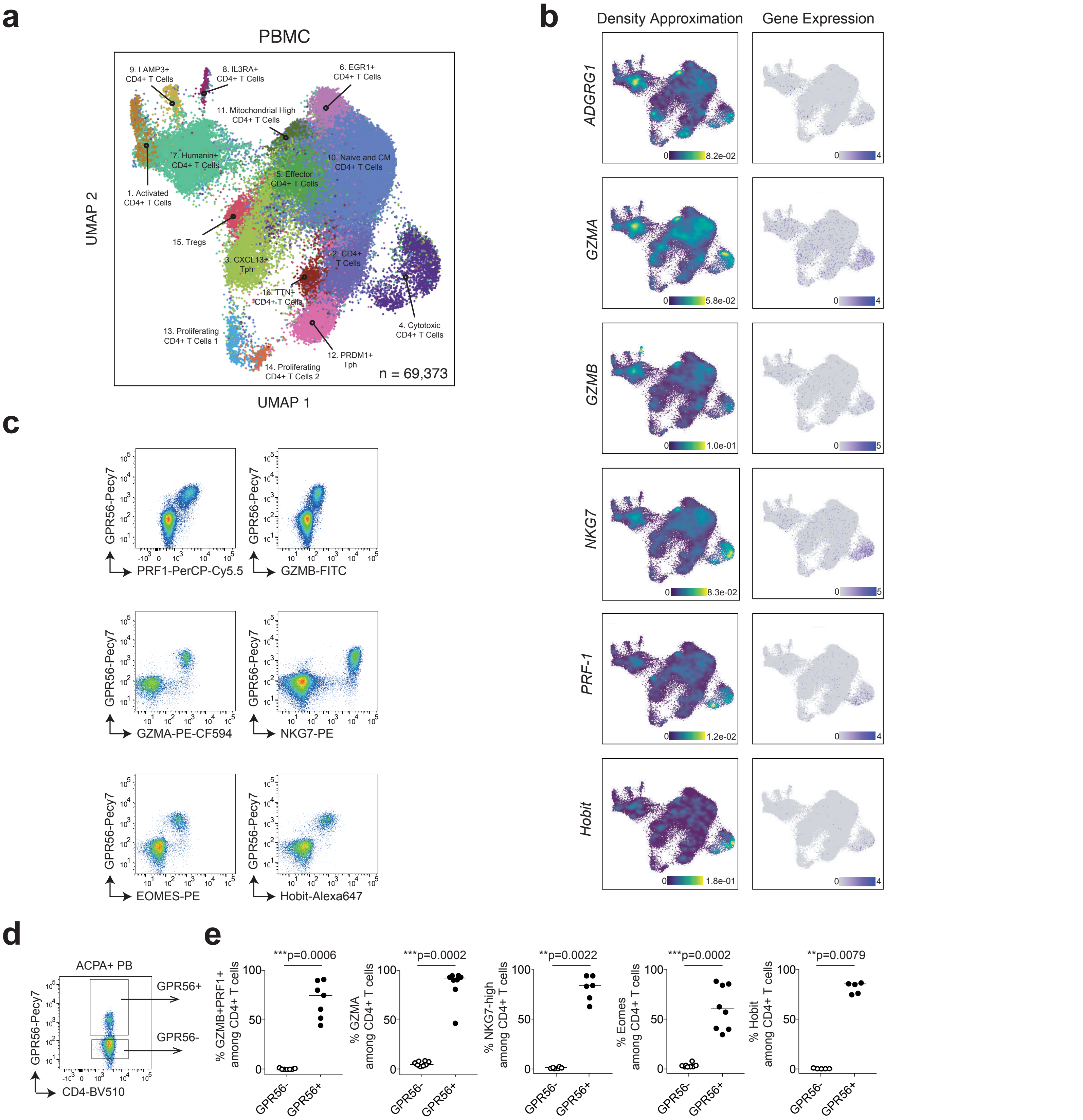


**Supplementary Fig. 5. Cytotoxic CD4+ T cells and GPR56 expression in PB from ACPA+ RA patients.** **a**) UMAP displaying 16 CD4+ T-cell clusters in PB in 7 ACPA+ RA patients. (**b**) Nebulosa density plots showing cytotoxic effector molecules and transcription factors in PB from 7 ACPA+ RA patients. (**c**) Representative flow cytometry dot plots showing the expression of effector molecules, receptors and transcription factors associated with cytotoxic functions in CD4+ T cells in ACPA+ RA PB (**d-e**) Frequency of GZMB, GZMA, NKG7, Eomes and Hobit in GPR56-negative and positive CD4+ T cells in ACPA+ RA PB (n=5-8). Line represents median, Mann-Whitney U test. Data are from a pool of seven independent experiments where a circle is a single replicate. White dots indicate GPR56- CD4+ T cells and black dots indicate GPR56+ CD4+ T cells.

### Supplementary Fig. 6

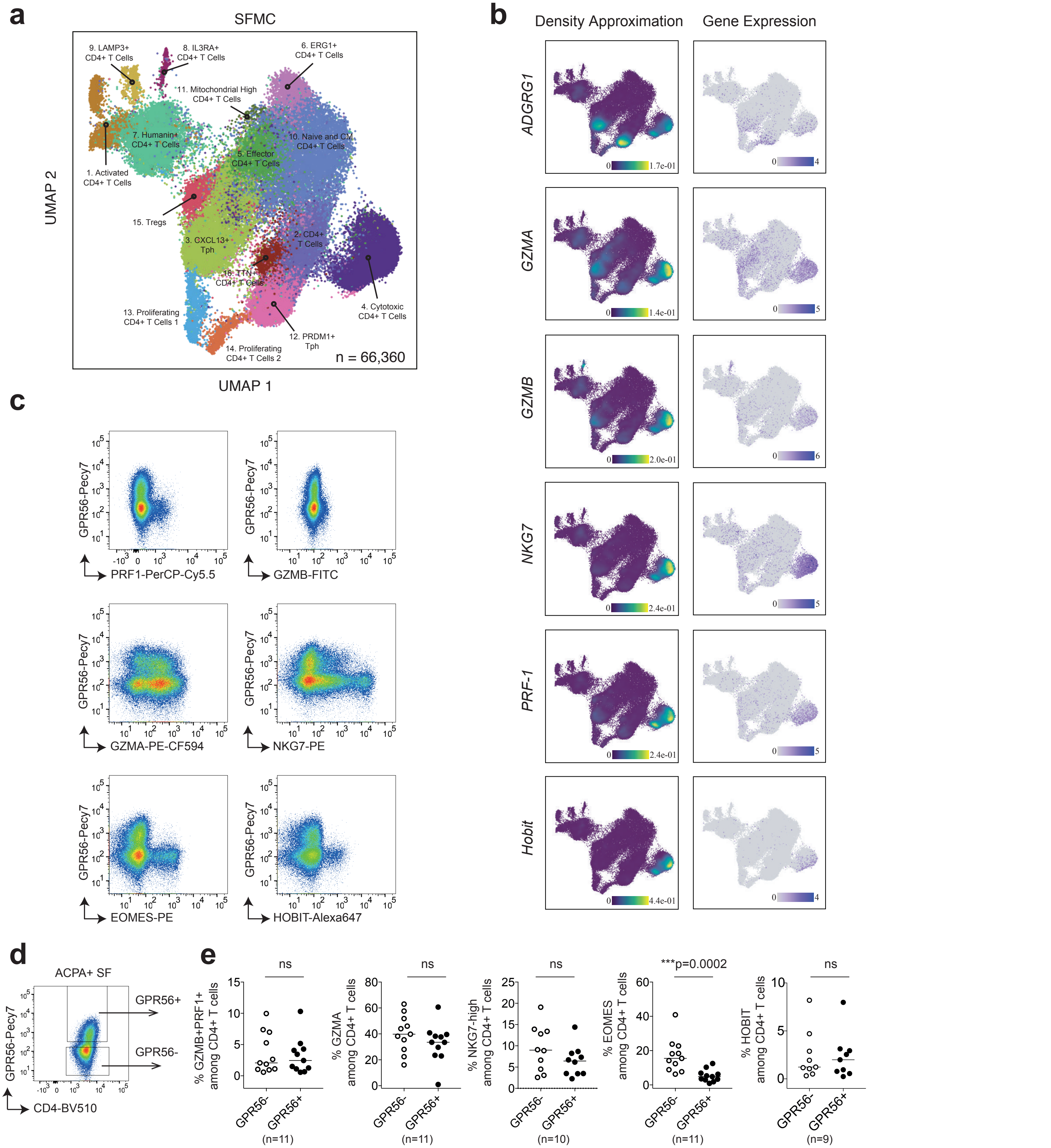


**Supplementary Fig. 6. Cytotoxic CD4+ T cells and GPR56 expression in SF from ACPA+ RA patients.** **a**) UMAP displaying 16 CD4+ T-cell clusters in PB in 7 ACPA+ RA patients. (**b**) Nebulosa density plots showing cytotoxic effector molecules and transcription factors in SF from 7 ACPA+ RA patients. (**c**) Representative flow cytometry dot plots showing the expression of effector molecules, receptors and transcription factors associated with cytotoxic functions in CD4+ T cells in SF ACPA+ RA SF (**d-e**) Frequency of GZMB, GZMA, NKG7, Eomes and Hobit in GPR56-negative and positive CD4+ T cells in ACPA+ RA SF (n=9-11). Line represents median, Mann-Whitney U test. Data are from a pool of nine independent experiments where a circle is a single replicate. White dots indicate GPR56- CD4+ T cells and black dots indicate GPR56+ CD4+ T cells.

### Supplementary Fig. 7

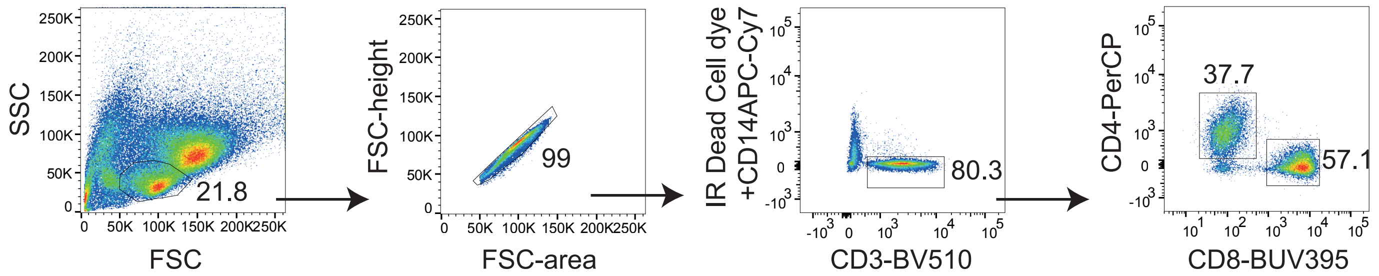


**Supplementary Fig. 7**. Facs gating strategy for Figure 4 and supp Fig 8 included lymphocyte gating (based on forward and side scatter) followed by doublets, CD14+ and dead cells exclusion. CD4+ or CD8+ T cells were further selected among CD3+ T cells.

### Supplementary Fig. 8

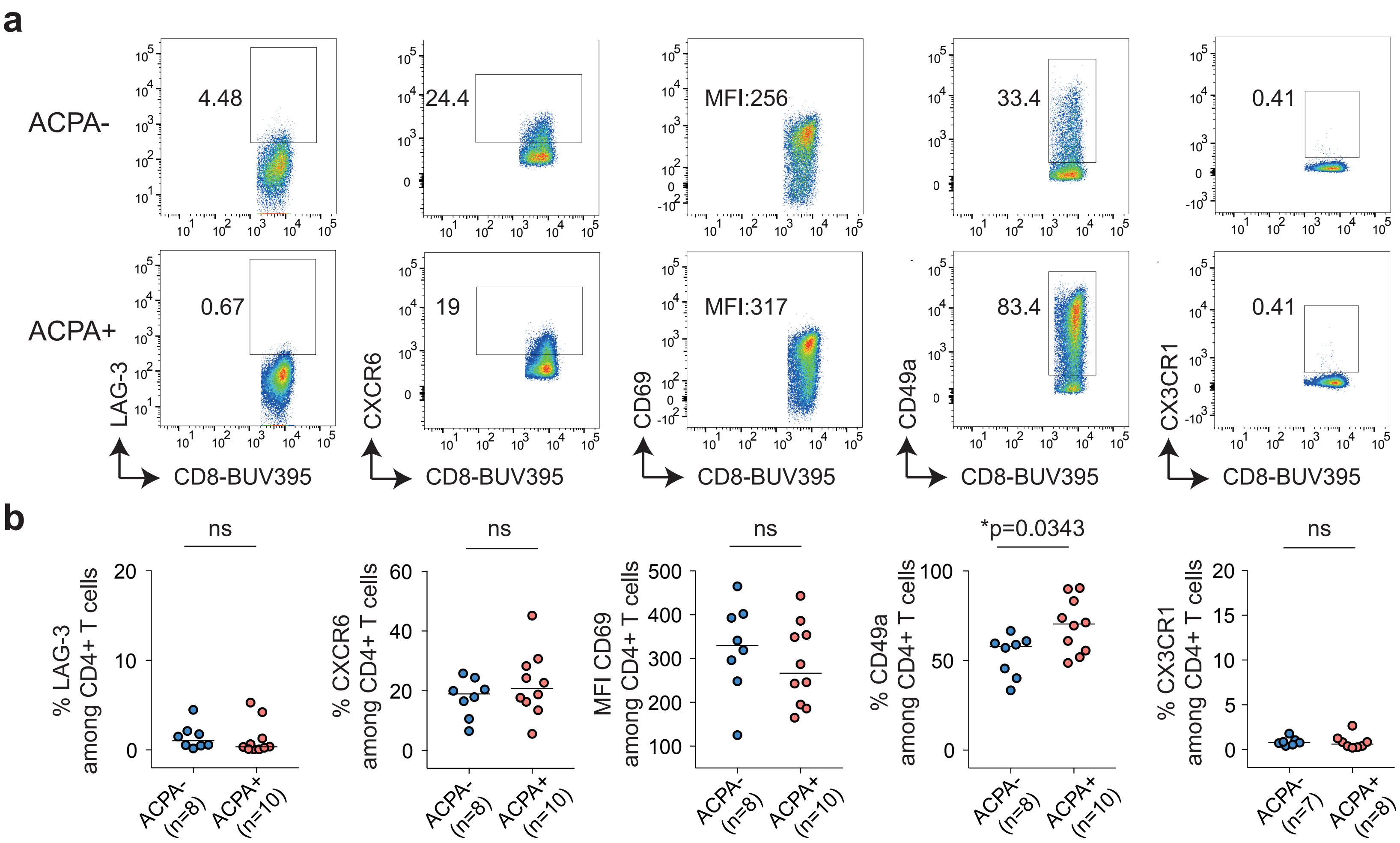


**Supplementary Fig. 8. Tissue-resident memory markers on CD8+ T cells in SF from RA patients**. **a**) Representative flow cytometry dot plots showing the expression of LAG-3, CXCR6, CD69, CD49a and CX3CR1 in ACPA- and ACPA+ SF CD8+ T cells, quantified in (**b**) (ACPA- n=7-8; ACPA+ n=8-10). Line represents median, Mann-Whitney U test. Data are from a pool of eight independent experiments where a circle is a single replicate. Blue dots indicate ACPA- RA SF and red dots indicate ACPA+ RA SF.

### Supplementary Fig. 9

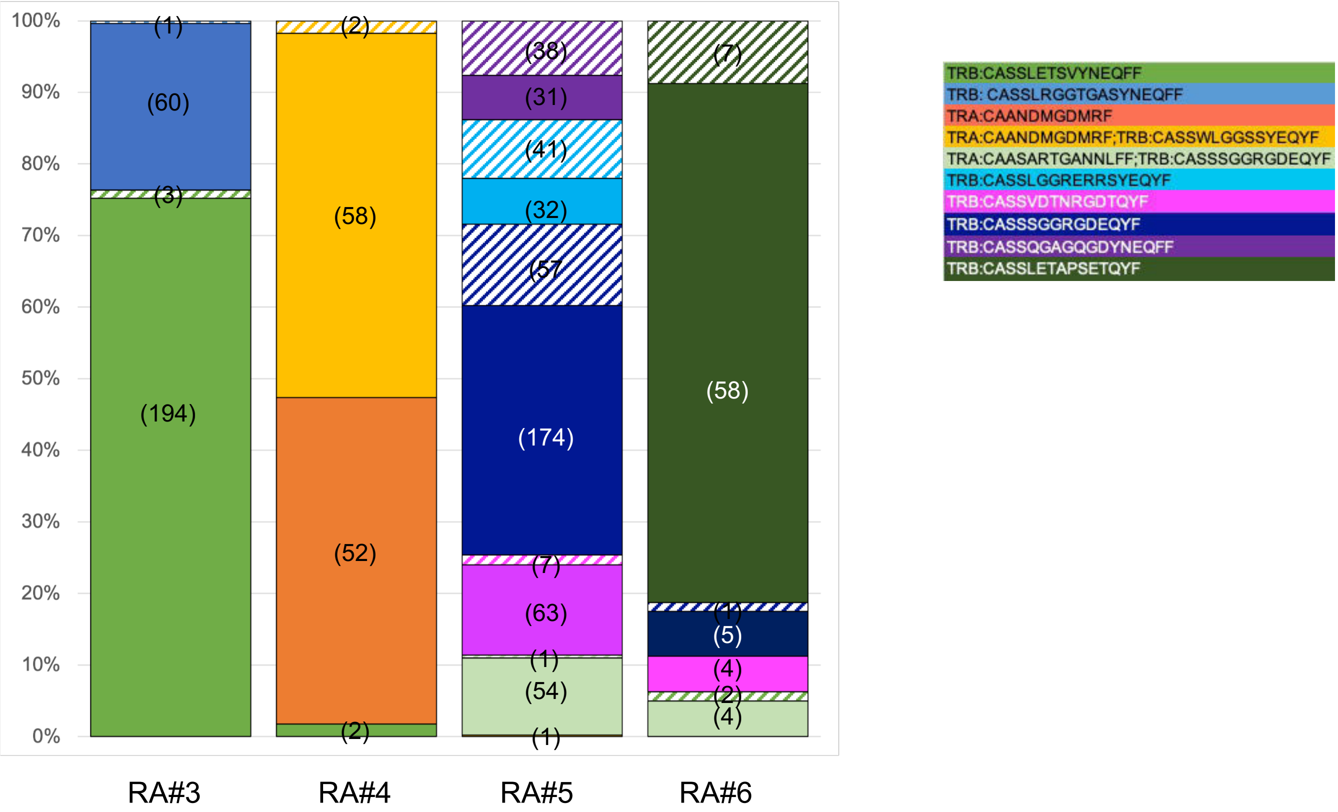


**Supplementary Fig 9. Top 10 expanded CD4⁺ T-cell clones.** Stacked barplots displaying the frequency of the top 10 expanded CD4+ T-cell clones in ACPA+ RA patients. Striped quadrants clones are from PBMC, full quadrants clones are from SFMC. Each color represents a clone. Numbers in parenthesis indicate absolute numbers.

**Supplementary Table 1**.Patients characteristics

| RA nr | Age[a] | Sex | anti-CCP titers* | RF | Disease duration | Methotrexate | Biologics | Cortocosteroids (dose) | Other synthetic DMARDS | Shared Epitope | HLA-DRB1 | 10X single cell sequencing PB and SF | Flow cytometry SF | Flow cytometry PB |
|---|---|---|---|---|---|---|---|---|---|---|---|---|---|---|
| RA1 | 60-65 | F | 890 | + | 6 | Y | N | N | N | N | *09/*09 | Y | Y | |
| RA2 | 60-65 | M | 1620 | + | 21 | N | abatacept (last infusion 4 months before) | Prednisolone 5mg | N | Y | *0101/*09 | Y | Y | Y |
| RA3 | 50-55 | F | 1360 | + | 24 | N | anti-CD20 (4 months before) | Prednisolone 10mg | N | N | *07/*15 | Y | Y | |
| RA4 | 55-60 | M | 790 | + | md | Y | N | N | N | md | md | Y | Y | Y |
| RA5 | 70-75 | F | 610 | md | 53 | N (stopped 3 months before) | N (abatacept discontinued 3 months before) | N | N | Y | *09/*10 | Y | Y | Y |
| RA6 | 35-40 | F | 1160 | - | 7 | N (stopped 3 months before) | anti-TNF | Prednisolone 5mg | N | N | *07/*09 | Y | Y | Y |
| RA7 | 60-65 | F | 160 | + | 43 | N | N | Prednisolone 5mg | N | Y | *04/*15 | Y | Y | |
| RA8 | 65-70 | F | 1780 | - | 14 | Y | N | Prednisolone 2.5mg | N | Y | *01/*14 | | Y | Y |
| RA9 | 70-75 | F | 4200 | + | 29 | N | abatacept (last infusion 2 weeks before) | N | N | Y | *03/*0401 | | Y | Y |
| RA10 | 60-65 | F | 1510 | + | 1 | N | N | Prednisolone 10mg | N | Y | *01/*14 | | Y | |
| RA11 | 70-75 | F | 580 | + | 2 | N (stopped 4 weeks before) | N | Prednisolone 10mg | N | N | *07/*13 | | Y | Y |
| RA12 | 45-50 | F | 210 | + | 25 | Y | anti-TNF | N | N | Y | *03/*0404 | | Y | Y |
| RA13 | 35-40 | F | neg | - | 12 | Y | N | N | Y | md | md | | Y | Y |
| RA14 | 50-55 | F | neg | - | 22 | Y | anti-TNF | N | N | Y | *01/*07 | | Y | |
| RA15 | 55-60 | M | neg | - | 6 | N | N | Prednisolone 10mg (stopped 2 weeks before) | N | Y | *0404/*15 | | Y | Y |
| RA16 | 60-65 | F | neg | - | 23 | Y | anti-TNF | Prednisolone 7.5mg | N | N | *0406/*11 | | Y | Y |
| RA17 | 45-50 | F | neg | - | 26 | N | abatacept (last infusion 2 weeks before) | Prednisolone 5mg | N | Y | *0101/*08 | | Y | Y |
| RA18 | 60-65 | F | neg | + | 3 | N | anti-IL-6R | Prednisolone 5mg | N | N | *03/*13 | | Y | Y |
| RA19 | 55-60 | M | neg | - | 29 | Y | anti-interleukin-17A | Prednisolone 10mg | N | Y | *01/*03 | | Y | Y |
| RA20 | 45-50 | M | neg | - | 21 | Y | N (anti-TNF stopped 4 months before) | No | N | Y | *01/*03 | | Y | Y |
| RA21 | 60-65 | F | neg | - | 4 | N | anti-IL-6R | Prednisolone 5 mg | N | N | *11/*13 | | Y | Y |

*cut-off for CCP = 25AU/ml
md: missing data
CCP: cyclic citrullinated peptide
RF: rheumatoid factor

**Supplementary Table 2**. Cell type composition (Frequencies)

PBMC

| RA patients | RA1 | RA2 | RA3 | RA4 | RA5 | RA6 | RA7 | Median |
|---|---|---|---|---|---|---|---|---|
| 1. Activated CD4+ T Cells | 0,62 | 0,63 | 0,61 | 0,42 | 5,43 | 0,48 | 0,96 | 0,62 |
| 2. CD4+ T Cells | 6,76 | 6,25 | 8,35 | 6,50 | 7,21 | 7,39 | 6,72 | 6,76 |
| 3. CXCL13+ Tph | 6,38 | 10,69 | 11,65 | 7,13 | 3,83 | 3,71 | 9,59 | 7,13 |
| 4. Cytotoxic CD4+ T Cells | 2,00 | 2,82 | 10,72 | 3,75 | 1,95 | 1,78 | 2,38 | 2,38 |
| 5. Effector CD4+ T Cells | 11,68 | 10,88 | 12,26 | 15,74 | 5,14 | 6,66 | 14,56 | 11,68 |
| 6. EGR1+ CD4+ T Cells | 2,44 | 1,88 | 3,02 | 3,85 | 1,74 | 22,94 | 2,22 | 2,44 |
| 7. Humanin+ CD4+ T Cells | 5,16 | 20,78 | 4,32 | 7,05 | 28,06 | 6,11 | 8,21 | 7,05 |
| 8. IL3RA+ CD4+ T Cells | 0,20 | 0,21 | 0,17 | 0,24 | 0,92 | 0,29 | 0,17 | 0,21 |
| 9. LAMP3+ CD4+ T Cells | 0,37 | 0,89 | 0,72 | 0,28 | 1,25 | 0,24 | 0,44 | 0,44 |
| 10. Naïve and CM CD4+ T Cells | 58,83 | 27,92 | 35,84 | 48,62 | 34,16 | 46,49 | 44,08 | 44,08 |
| 11. Mitochondrial High CD4+ T Cells | 0,68 | 9,85 | 2,68 | 1,52 | 1,47 | 0,15 | 0,13 | 1,47 |
| 12. PRDM1+ Tph | 1,92 | 3,00 | 3,61 | 1,51 | 4,55 | 1,93 | 4,00 | 3,00 |
| 13. Proliferating CD4+ T Cells 1 | 0,32 | 1,55 | 1,31 | 0,46 | 1,47 | 0,20 | 1,07 | 1,07 |
| 14. Proflierating CD4+ T Cells 2 | 0,25 | 0,20 | 0,40 | 0,20 | 0,65 | 0,23 | 0,57 | 0,25 |
| 15. Tregs | 1,65 | 1,22 | 2,41 | 1,93 | 0,93 | 0,89 | 3,63 | 1,65 |
| 16. TTN+ CD4+ T Cells | 0,74 | 1,25 | 1,94 | 0,80 | 1,25 | 0,50 | 1,26 | 1,25 |

SFMC

| RA patients | RA1 | RA2 | RA3 | RA4 | RA5 | RA6 | RA7 | Median |
|---|---|---|---|---|---|---|---|---|
| 1. Activated CD4+ T Cells | 0,19 | 7,43 | 3,62 | 2,82 | 1,80 | 1,83 | 4,52 | 2,82 |
| 2. CD4+ T Cells | 18,75 | 7,27 | 3,72 | 6,64 | 6,34 | 8,20 | 16,73 | 7,27 |
| 3. CXCL13+ Tph | 13,25 | 21,36 | 8,40 | 27,61 | 15,27 | 13,13 | 23,50 | 15,27 |
| 4. Cytotoxic CD4+ T Cells | 11,81 | 3,10 | 55,05 | 12,63 | 8,77 | 1,86 | 3,51 | 8,77 |
| 5. Effector CD4+ T Cells | 14,60 | 1,51 | 2,45 | 11,42 | 5,40 | 8,76 | 5,01 | 5,40 |
| 6. EGR1+ CD4+ T Cells | 2,27 | 0,51 | 0,48 | 1,00 | 1,09 | 16,58 | 1,18 | 1,09 |
| 7. Humanin+ CD4+ T Cells | 4,87 | 18,40 | 4,97 | 3,39 | 34,72 | 2,89 | 9,09 | 4,97 |
| 8. IL3RA+ CD4+ T Cells | 0,00 | 0,42 | 0,41 | 0,30 | 0,80 | 0,39 | 1,20 | 0,41 |
| 9. LAMP3+ CD4+ T Cells | 0,14 | 1,04 | 1,05 | 1,02 | 0,51 | 1,25 | 0,83 | 1,02 |
| 10. Naïve and CM CD4+ T Cells | 11,04 | 5,49 | 4,96 | 8,33 | 9,30 | 32,48 | 17,91 | 9,30 |
| 11. Mitochondrial High CD4+ T Cells | 0,19 | 0,36 | 0,24 | 0,46 | 0,10 | 0,15 | 0,11 | 0,19 |
| 12. PRDM1+ Tph | 8,39 | 22,72 | 7,00 | 11,07 | 7,30 | 8,69 | 6,97 | 8,39 |
| 13. Proliferating CD4+ T Cells 1 | 6,41 | 2,44 | 1,53 | 4,76 | 1,60 | 1,63 | 0,95 | 1,63 |
| 14. Proflierating CD4+ T Cells 2 | 3,23 | 2,30 | 1,42 | 2,45 | 1,07 | 0,82 | 0,73 | 1,42 |
| 15. Tregs | 2,31 | 2,73 | 2,21 | 4,29 | 3,70 | 0,82 | 4,85 | 2,73 |
| 16. TTN+ CD4+ T Cells | 2,55 | 2,92 | 2,48 | 1,78 | 2,24 | 0,52 | 2,89 | 2,48 |

**Supplementary Table 3**. Flow cytometry antibodies

| Antibody | Fluorochrome | Clone | Source |
|---|---|---|---|
| CD14 | APC-H7 | MφP-9 | BD Biosciences |
| CD3 | BV786 | UCHT1 | BD Biosciences |
| CD3 | BV510 | UCHT-1 | BD Biosciences |
| CD4 | BV510 | SK3 | BD Biosciences |
| CD4 | PerCP | RPA-T4 | Biolegend |
| CD8 | BUV395 | RPA-T8 | BD Biosciences |
| PD-1 | BV421 | EH12.1 | BD Biosciences |
| PD-1 | BB515 | EH12.1 | BD Biosciences |
| HLA-DR | BV711 | G46-6 | BD Biosciences |
| GPR56 | PE-Cy7 | CG4 | Biolegend |
| CXCR6 | Alexa Fluor 647 | K041E5 | Biolegend |
| LAG3 | BV421 | 11C3C65 | Biolegend |
| CD49a | PE | TS2/7 | Biolegend |
| CD69 | BV786 | FN50 | BD Biosciences |
| CX3CR1 | FITC | 2A9-1 | Biolegend |
| GZMB | FITC | GB11 | BD Biosciences |
| GZMA | Alexa Fluor 594 | CB9 | Biolegend |
| PRF1 | PercP/Cyanine5.5 | B-D48 | Biolegend |
| Hobit | Alexa Fluor 647 | Sanquin-Hobit/1 | BD Biosciences |
| NKG7 | PE | 2G9 | Beckman Coulter |
| Eomes | PE | WD1928 | eBioscience |

**Supplementary Table 4**. Flow cytometry panels

| Panel 1 | Fluorochrome | Clone | Source |
|---|---|---|---|
| Viability dye | Near infra-red | L34975 | ThermoFischer Scientific |
| CD14 | APC-H7 | MφP-9 | BD Biosciences |
| CD3 | BV786 | UCHT1 | BD Biosciences |
| CD4 | BV510 | SK3 | BD Biosciences |
| CD8 | BUV395 | RPA-T8 | BD Biosciences |
| PD-1 | BV421 | EH12.1 | BD Biosciences |
| HLA-DR | BV711 | G46-6 | BD Biosciences |
| GPR56 | PE-Cy7 | CG4 | Biolegend |
| GZMA | Alexa Fluor 594 | CB9 | Biolegend |
| PRF1 | PercP/Cyanine5.5 | B-D48 | Biolegend |
| Hobit | Alexa Fluor 647 | Sanquin-Hobit/1 | BD Biosciences |
| Eomes | PE | WD1928 | eBioscience |

| Panel 2 | Fluorochrome | Clone | Source |
|---|---|---|---|
| Viability dye | Near infra-red | L34975 | ThermoFischer Scientific |
| CD14 | APC-H7 | MφP-9 | BD Biosciences |
| CD3 | BV786 | UCHT1 | BD Biosciences |
| CD4 | BV510 | SK3 | BD Biosciences |
| CD8 | BUV395 | RPA-T8 | BD Biosciences |
| PD-1 | BV421 | EH12.1 | BD Biosciences |
| GPR56 | PE-Cy7 | CG4 | Biolegend |
| GZMB | FITC | GB11 | BD Biosciences |
| GZMA | Alexa Fluor 594 | CB9 | Biolegend |
| PRF1 | PercP/Cyanine5.5 | B-D48 | Biolegend |
| Hobit | Alexa Fluor 647 | Sanquin-Hobit/1 | BD Biosciences |
| NKG7 | PE | 2G9 | Beckman Coulter |

| Panel 3 | Fluorochrome | Clone | Source |
|---|---|---|---|
| Viability dye | Near infra-red | L34975 | ThermoFischer Scientific |
| CD14 | APC-H7 | MφP-9 | BD Biosciences |
| CD3 | BV510 | UCHT-1 | BD Biosciences |
| CD4 | PerCP | RPA-T4 | Biolegend |
| CD8 | BUV395 | RPA-T8 | BD Biosciences |
| PD-1 | BB515 | EH12.1 | BD Biosciences |
| HLA-DR | BV711 | G46-6 | BD Biosciences |
| GPR56 | PE-Cy7 | CG4 | Biolegend |
| CXCR6 | Alexa Fluor 647 | K041E5 | Biolegend |
| LAG3 | BV421 | 11C3C65 | Biolegend |
| CD49a | PE | TS2/7 | Biolegend |
| CD69 | BV786 | FN50 | BD Biosciences |

| Panel 4 | Fluorochrome | Clone | Source |
|---|---|---|---|
| Viability dye | Near infra-red | L34975 | ThermoFischer Scientific |
| CD14 | APC-H7 | MφP-9 | BD Biosciences |
| CD3 | BV786 | UCHT1 | BD Biosciences |
| CD4 | BV510 | SK3 | BD Biosciences |
| CD8 | BUV395 | RPA-T8 | BD Biosciences |
| PD-1 | BV421 | EH12.1 | BD Biosciences |
| HLA-DR | BV711 | G46-6 | BD Biosciences |
| GPR56 | PE-Cy7 | CG4 | Biolegend |
| CXCR6 | Alexa Fluor 647 | K041E5 | Biolegend |
| CD49a | PE | TS2/7 | Biolegend |
| CX3CR1 | FITC | 2A9-1 | Biolegend |
